# Fecal Short-Chain Fatty Acids Vary by Sex and Amyloid Status

**DOI:** 10.1101/2025.07.31.25332523

**Authors:** Jessamine F. Kuehn, Qijun Zhang, Margo B. Heston, Jea Woo Kang, Sandra J. Harding, Nancy J. Davenport-Sis, Darby C. Peter, Robert L. Kerby, Vaibhav Vemuganti, Emma C. Schiffmann, Michael M. Tallon, Joseph Harpt, Arun Hajra, J. L. Wheeler, Sushma Shankar, Alissa Mickol, Justus Zemberi, Hana Chow, Eric Zhang, Eleanor Clements, Hanna Noughani, Allison Forst, Grace Everitt, Gwendlyn Kollmorgen, Clara Quijano-Rubio, Bradley T. Christian, Cynthia M. Carlsson, Sterling C. Johnson, Sanjay Asthana, Henrik Zetterberg, Kaj Blennow, Tyler K. Ulland, Federico E. Rey, Barbara B. Bendlin

## Abstract

**INTRODUCTION:** Short-chain fatty acids (SCFAs), produced by gut microbes, influence Alzheimer’s disease (AD) pathology in animals. Less is known about SCFAs and AD in humans. We profiled feces of adults along the AD continuum to investigate gut microbiome and SCFA associations with AD pathology and cognition.

**METHODS:** We measured SCFAs and bacterial abundances in fecal samples from 287 participants in the Wisconsin Alzheimer’s Disease Research Center and Wisconsin Registry for Alzheimer’s Prevention. We performed regressions examining associations between SCFAs or gut microbes and AD pathology and cognition.

**RESULTS:** Fecal propionate, isovalerate, and propionate-producing bacteria are inversely associated with amyloid status. Mediation analysis found that propionate mediates sex-specific associations between SCFAs and CSF biomarkers. SCFA levels are associated with slower cognitive decline.

**DISCUSSION:** These results link SCFAs and propionate-producing bacteria with AD. This may inform efforts to leverage diet and specific bacteria to boost SCFA production and potentially ameliorate AD progression.

## 1. Background

With advances in Alzheimer’s disease (AD) therapeutics and findings that suggest AD pathology may be influenced by lifestyle and environmental factors, there is a mounting need to identify biological mechanisms that underlie modifiable risk factors. Core features of AD pathogenesis include the accumulation of extracellular amyloid plaques and intraneuronal neurofibrillary tau tangles in the central nervous system, in tandem with glial activation, followed by neurodegeneration and amnestic cognitive decline[1]. Among several proposed modifiable risk factors, there is increasing evidence suggesting gut-brain axis involvement in AD pathology. Human studies have noted gut microbiome changes in older adults with mild cognitive impairment or dementia[2–4], and have identified microbial associations with amyloid positron emission tomography (PET) neuroimaging and fluid biomarkers indicative of amyloid, tau, and neuroinflammation and neurodegeneration[5]. Animal studies have also noted that AD mouse models harbor different gut microbiomes compared to wild-type (WT) mice[6], and that altering their microbiomes can result in changes to AD pathology[7]. For instance, in the APP/PS1 mouse model of amyloid beta (Aβ) plaque accumulation, mice raised in sterile environments without gut microbiota have reduced amyloid burden compared to conventionally-raised counterparts[8]. Furthermore, APP/PS1 mice given fecal microbiota transplants from WT mice have ameliorated AD pathology compared to untreated APP/PS1 mice[9]. Although mounting evidence suggests a potential role of the gut microbiome in AD, the mechanisms by which microbes influence disease progression remain unclear.

The aging gut microbiome may increase risk for AD by contributing to intestinal inflammation[10]. Short-chain fatty acids (SCFAs) are produced in the colon via bacterial fermentation of dietary substrates, especially plant polysaccharides, that reach the distal gut[11]. The role of SCFAs in health and disease is complex, however SCFAs are generally associated with host health; for example, they promote gut barrier integrity and reduce inflammation[12,13]. SCFA levels are reduced in patients with intestinal, immune, and neurological diseases[14–16], several of which are associated with AD[10,17]. Decreased SCFA levels have been observed in AD mice[18], and increases in SCFA levels can have a beneficial effect on AD pathology in mice: one study of 3xTg mice found that supplementation of tributyrin, a prodrug of butyrate, prevented tau hyperphosphorylation and abrogated memory deficits[19]. In humans, one small study (n=17) showed that diet alterations could affect amyloid and tau pathology through both shifting microbiome composition and SCFA production[4]. Although SCFAs may be implicated in mechanisms of brain health and have been found to improve AD pathology in mice, and the gut microbiome and intestinal health are associated with AD in humans, there is currently no direct evidence in a large preclinical cohort that connects microbiome composition, SCFAs, and AD pathological and cognitive outcomes in humans.

Given the well-established anti-inflammatory properties of SCFAs, the current study builds on our prior finding that intestinal inflammation is elevated in aging and associated with AD pathology[10]. We performed a multimodal analysis in humans to investigate SCFA relationships with AD biomarkers and cognitive decline, and used mediation analysis to determine the role of the gut microbiome in these relationships. The goals of this study were to 1) Characterize SCFA relationships with amyloid and tau status, cerebrospinal fluid (CSF) biomarkers of AD, neuroinflammation, and neurodegeneration, and trajectories of plasma phosphorylated tau (pTau)_217_ and cognitive performance, 2) Determine the extent to which abundances of bacteria with genes encoding production of the SCFAs propionate and butyrate are related to these biomarkers, and 3) Test the extent to which SCFA abundance mediates relationships between SCFA-producing bacteria and biomarkers. Given prior evidence suggesting sex differences in both gut microbiome composition[20,21] and AD trajectories[22,23], we also tested SCFA-by-sex and microbiome-by-sex interaction effects.

## 2. Methods

### 2.1. Participant inclusion criteria

Participants were recruited into the Microbiome and Alzheimer’s Risk Study (MARS) from the Wisconsin Alzheimer’s Disease Research Center (ADRC) and Wisconsin Registry for Alzheimer’s Prevention (WRAP), two longitudinal prospective cohort studies of midlife and older adults that are enriched for risk of late-onset AD due to parental history of AD[24]. Exclusion criteria for the Wisconsin ADRC and WRAP studies include any significant neurologic disease other than AD, history of alcohol/substance dependence, major psychiatric disorders, or other significant medical illness. Participants in the MARS study were required to be at least 40 years of age, in good general health (other than dementia), and commit to fecal sample collection. Participants in the present analysis also had previous or current enrollment in biomarker studies including the Wisconsin ADRC[25], WRAP[24], the Alzheimer’s Disease Connectome Project (ADCP)[26], or the Synapse Project (SV2A PET Imaging in Alzheimer’s Disease) studies. Prospective participants were excluded from MARS if they used immune stimulating medications aside from vaccines within 30 days, used methotrexate or immunosuppressive cytotoxic agents within 30 days, consumed antibiotics within 30 days, adopted a substantial dietary change (eliminating/significantly increasing major food group) within the last 30 days, or provided a sample with a Bristol Stool Score (BSS) of 7 (indicating a liquid stool consistency) since these samples have diminished species’ richness and altered gut microbiome composition[27]. For the current study, participants were included in biomarker and cognitive analyses if they had completed fecal sample collection and had biomarker and cognitive assessments proximal to the time of fecal collection, including amyloid or tau PET neuroimaging data within 5 years, and cognitive testing, CSF or plasma data within 2 years (**Fig. 1**). The cohort comprised a proportionally small group of participants with dementia; because we primarily studied preclinical associations with AD pathology and cognitive decline, we report results from the cognitively unimpaired (CU) subset here and results from the full cohort in the Supplementary Materials.

**Figure 1.**
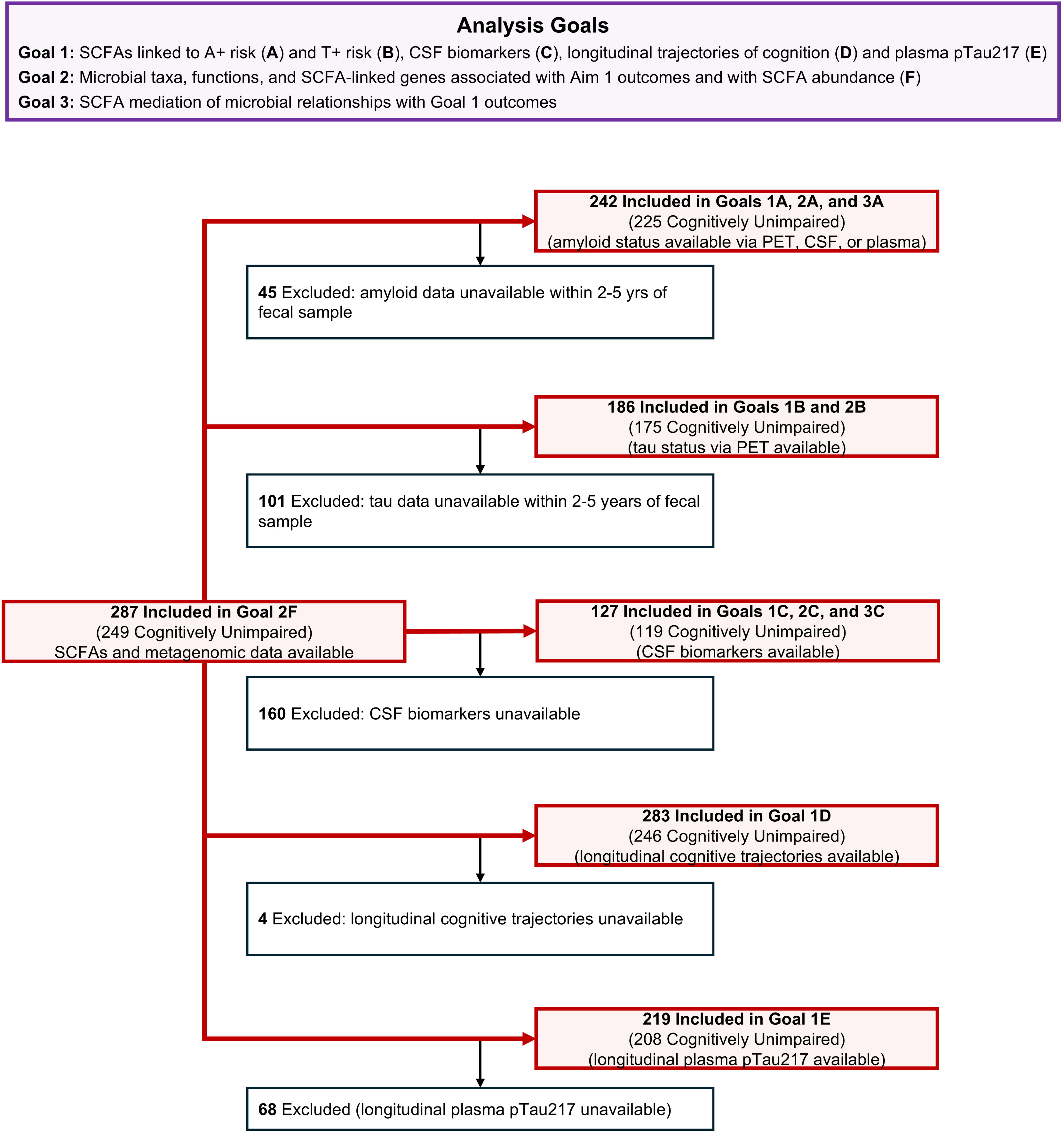
Analysis dataset selection per study goal. CONSORT flow diagram indicating selection criteria for inclusion in each analysis goal. Abbreviations: A+, amyloid status determined with ^11^C-PiB PET, CSF Aβ_42_/Aβ_40_, or plasma pTau_217_; *APOE*, apolipoprotein E; CSF, cerebrospinal fluid; PET, positron emission tomography; SCFA, short-chain fatty acids measured in fecal samples; T+, tau positive status measured with ^18^F-MK6240 PET neuroimaging.

All study procedures and analyses were approved by the University of Wisconsin (UW) Health Science Institutional Review Board, and all experiments were conducted in accordance with the Declaration of Helsinki and relevant guidelines and regulations (Wisconsin ADRC protocol ID: 2015-0030; WRAP protocol ID: 2023-1522; WRAP and Wisconsin ADRC umbrella protocol ID: 2013-0178; MARS protocol ID: 2015-1121; Synapse protocol ID: 2018-1283; ADCP protocol ID: 2022-0320). Prior to admission to the study, participants provided informed consent to be involved in the study.

### 2.2. Clinical measurements

Participants completed annual and biannual study visits during which physical, neurological, and neuropsychological exams were performed, and neuroimaging and fluid biosamples were collected. Clinical diagnoses of cognitively unimpaired, mild cognitive impairment, or dementia were based on the NINDS/ADRDA criteria[28,29] and confirmed by a multidisciplinary expert panel. For the current study, clinical outcomes included cognitive trajectories defined with a Preclinical Alzheimer’s Cognitive Composite (PACC3)[30], which was derived by averaging the standardized z-scored Rey Auditory Verbal Learning Test (RAVLT) summed learning trials, Logical Memory II Story A, and Trail-Making Test Part B (TMT-B) scores. Clinical covariates in statistical models included *APOE* ε4 carriage determined with blood samples[31] and body mass index (BMI). Data on medication and dietary supplement usage were also collected at each visit. To rule out the possibility that medications or supplements could influence relationships observed between biomarker outcomes and microbiome composition or SCFA levels, we compared usage rates of each class of medication or supplement across diagnosis and amyloid status, defining usage as a binary variable (using vs. not using) (Supplementary Table 1).

### 2.3. AD biomarker measurement

Biomarker outcomes were obtained through serial study visits at UW-Madison. Neuroimaging scans were acquired at the Waisman Center and the Wisconsin Institute for Medical Research, and processing and quantification were performed locally. CSF and blood samples were acquired at the Clinical Research Unit within the UW Hospital; samples were processed and stored locally at -80°C. CSF biomarkers were measured by the Clinical Neurochemistry Laboratory at the University of Gothenburg, and pTau_217_ in blood plasma was measured by the Wisconsin ADRC Biofluid Laboratory. Biomarker outcomes included 1) amyloid-positive (A+) or negative (A-) status, 2) tau positive (T+) or negative (T-) status, 3) CSF biomarkers of AD pathology, glial activation, and neurodegeneration measured continuously, and 4) continuous trajectories of plasma pTau_217_. A+/- status was determined using ^11^C-Pittsburgh compound B (^11^C-PiB) PET neuroimaging obtained within five years of fecal collection, or CSF Aβ_42/40_ or plasma pTau_217_ obtained within two years of fecal collection. For participants with more than one amyloid biomarker measured, the biomarker time point closest to fecal collection was used as the indicator of amyloid status. T+/- status was determined using ^18^F-MK620 PET neuroimaging obtained within five years of fecal collection.

#### 2.3.1. Amyloid and tau PET acquisition, processing and quantification

^11^C-PiB and ^18^F-MK6240 PET scans were acquired and quantified using previously published methods[32,33] at the University of Wisconsin-Madison (UW-Madison) Waisman Brain Imaging Lab to measure cortical amyloid and tau burden, respectively. For both amyloid and tau PET, images were processed with an MRI-guided pipeline using SPM12. Logan graphical analysis[34] was used to generate ^11^C-PiB distribution volume ratios (DVRs) in subject space, normalized by a cerebellar gray matter reference region (MICCAI cerebellum atlas)[35]. Global cortical mean DVR was calculated per scan by averaging DVR across eight bilateral gray matter regions of interest defined with the Automated Anatomical Labelling Atlas(AAL)-3 (anterior cingulate cortex, angular gyrus, middle temporal gyrus, posterior cingulate cortex, precuneus, supramarginal gyrus, superior temporal gyrus, ventromedial prefrontal cortex)[36]. Participants were considered A+ using a previously defined threshold of global cortical DVR>1.16. ^18^F-MK6240 data obtained 70-90 minutes post-injection were used to generate standard uptake value ratios (SUVRs) in subject space. The 5-minute time frames were first realigned, summed, coregistered to the T1-weighted MRI and spatially normalized into standardized space by inferior cerebellar gray matter defined using an eroded AAL-3 mask. A meta-temporal ROI was applied to the PET image and divided by the voxel average of the inferior cerebellar grey matter reference region to obtain the SUVR measure. Participants were classified as T+ using a previously defined threshold of entorhinal SUVR>1.27, representing elevated tau burden in the first Braak neuropathological staging region for neurofibrillary tangles[37].

#### 2.3.2. CSF biomarker measurement

127 of the participants included in this study underwent lumbar puncture for CSF collection. Most participants fasted for 9-12 hours before CSF collection in the morning as previously described (N=3 did not fast)[38]. CSF was collected using the Wisconsin ADRC CSF biomarker service procedures[38]. CSF biomarker levels for Aβ42, pTau181, and tTau were measured using the Elecsys® β-Amyloid (1–42) CSF I and II, Elecsys Phospho-Tau (181P) CSF and Elecsys Total-Tau CSF assays on the Cobas® e 601 analyzer (all Roche Diagnostics International Ltd, Rotkreuz, Switzerland). CSF biomarker levels for Aβ40, axonal degeneration (neurofilament light protein[NfL]), synaptic degeneration (neurogranin, α-synuclein), reactive astrocytes (glial fibrillary acidic protein[GFAP]), glial activation (chitinase-3-like protein 1[YKL-40]), and microglial activation (soluble triggering receptor expressed on myeloid cells 2[sTREM2]) were measured using the NeuroToolKit, a panel of exploratory robust prototype assays, on the Cobas e 411 analyzer (both Roche Diagnostics International Ltd) [30, 42]. Calculations of Aβ42/Aβ40 and pTau181/Aβ42 ratios were made using the biomarker results. Assays were completed in two batches; batch 2 included the Elecsys® β-Amyloid (1–42) CSF II assay that has superior detection limits. Based on A+ thresholds calculated with receiver operator characteristic analysis[38], participants were determined to be A+ at Aβ_42_/Aβ_40_<0.046 and Aβ_42_/Aβ_40_<0.055 for batch 1 and 2, respectively. In statistical analyses featuring Aβ_42_/Aβ_40_ as a continuous outcome, we controlled for inter-batch variability by including batch number as a covariate.

#### 2.3.3. Plasma biomarker measurement

Blood was collected under 12-hour fasting conditions between August 2011 and July 2023 into tubes containing heparin and K_2_ EDTA from 2011 to August 2020 and September 2020 to 2023, respectively. After collection, plasma samples were inverted, centrifuged, divided into aliquots, and stored at -80^°^C. Preanalytic protocol details between 2011 and 2023 are shown in Supplementary Table 2. The ALZpath pTau_217_ version 2 assay was used across the WRAP and Wisconsin ADRC cohorts to measure plasma levels of pTau_217_, a biomarker highly correlated with amyloid PET[39]. Assays were performed in duplicate on the Quanterix HD-X platform using two reagent kit lots. A bridging study used 35 samples to test the relationship between lots (Pearson correlation L=0.97, *p*<2.2e-16), and linear regression was applied to harmonize the data across reagent lots. We used previously published thresholds to indicate A-status at pTau_217_<0.4 pg/mL and A+ status at pTau_217_>0.63 pg/mL[39]. Plasma biomarkers were not used to indicate amyloid status for participants with pTau_217_ levels within the indeterminate range (0.4≤pTau_217_≤0.63 pg/mL, n=48).

### 2.4. Fecal Analysis

#### 2.4.1. Sample Collection

Between 2015 and 2023 participants collected fecal samples using collection kits at home and returned the samples to the University of Wisconsin-Madison Bacteriology Department using overnight delivery with frozen gel packs. Questionnaires describing participants’ medical history and diet in the past year accompanied the samples. Fecal samples were processed when received by lab members blinded to participant demographics. Each sample was weighed and scored with a BSS ranging from 1 to 7. Approximately 2-4 g of the sample was stored in two sterile straws in a 15 ml conical tube at −80°C using published methods[40]. All samples underwent the above preanalytical protocol except for the first samples collected for MARS (n=45 in the current study’s analysis cohort), which were stored in their collection tubs at −80°C.

#### 2.4.2. SCFA Quantification

Fecal SCFA levels were measured by headspace gas chromatography (GC) in a blinded manner. Samples from 287 participants were included. For each participant, a sterile fecal sample 50-300 mg was weighed, and milliQ water was added to adjust the total volume to 300 µL per sample. In a chilled 20 mL headspace GC septum-topped vial, 2.0 g (+/- 0.5 g) NaHSO_4_, the sample, milliQ water, and 1.0 mL of pre-chilled 60 µM 2-butanol (internal standard) was added. Vials were crimped and sat overnight for 1-3 days at room temperature. Samples were prepared in random order. For standards, a solution of acetate, propionate, butyrate, isobutyrate, valerate, and isovalerate (150 mM, 20 mM, 10 mM, 20 mM, 10 mM, 10 mM) was prepared, and from this seven serial 1:1 dilutions in milliQ water were generated. 300 µL of each dilution was prepared in a vial with NaSO_4_ and 2-butanol as described. These seven standards were prepared and included in each run and used to create standard curves for SCFA quantification (R^2^>0.97 for all standard curves).

SCFAs acetate, propionate, butyrate, isobutyrate, valerate, and isovalerate were measured using a Shimadzu GC-2010 Plus GC system (Shimadzu, Addison, IL) equipped with a flame ionization detector (FID). Vials were loaded onto the GC in a random order, and incubated at 80°C for 20 min before headspace sampling using a Shimadzu HS-20 headspace sampler (vials pressurized to 80 kPa with N_2_) and loading on a 2 mL sample loop heated to 150°C. The GC inlet was held at 150°C, and the inlet split ratio was set to 15:1. Samples were chromatographed on a PolarWax column (Restek, Belefonte, PA) (30 m × 0.25 mm, 0.10 μm film thickness) with N_2_ as carrier at a constant flow rate of 29.3 cm s−1. Column temperature was 40°C for 2 min then ramped to 200°C at 20 °C/min. LabSolutions software (Shimadzu, Addison, IL) was used to acquire and process data using peak areas and standards from known concentrations of the SCFAs. Measured SCFA levels are replicable across technical replicates (Supplementary Fig. 1) and across batches (Supplementary Fig. 2).

#### 2.4.3. DNA Extraction, Metagenomic Sequencing, and Pre-Analytic Processing

DNA was isolated from the fecal samples of the participants using a phenol:chloroform plus bead beating protocol for shotgun metagenomic deep sequencing with Illumina NovaSeq (∼32 million reads per sample). Low quality reads and host sequences were filtered out using Trimmomatic, bowtie2, and samtools (Supplementary Methods).

#### 2.4.4. Profiling microbiome composition, functional pathways, metagenome-assembled genomes (MAGs), and SCFA gene annotation

Reads were mapped to taxa and genetic pathways using the MetaPhlAn4[41] and humann3 databases[42]. Additionally, *de novo* metagenome assembly was used to identify metagenome-assembled genomes (MAGs) of microbial taxa, using SPAdes, Bowtie2, and MetaBAT2. 1464 high-quality MAGs were identified. Gene annotation with hidden Markov models using MUSCLE and HMMER was applied to identify putative genes encoding the production of butyrate and propionate, the two most abundant SCFAs in the gut that are exclusively produced by gut microbes[43,44]. MAGs were annotated as butyrate-producing genomes if they contained all four genes encoding for enzymes involved in the acetyl-CoA pathway (*thl, bhbd, cro, but*)[45,46]. MAGs were annotated as propionate-producing genomes if they contained all three genes encoding for enzymes in the succinate pathway (*mut, epi, mmd*) or both genes encoding for enzymes in the propanediol pathway (*PduCDE, PduP*)[45] (Supplementary Methods).

### 2.5 Statistical Analysis

All statistical analyses were performed using R software version 4.3.2 and packages stats (4.3.2), lavaan (0.6-17), mediation (4.5.0), vegan (2.6-10), dplyr (1.1.4), and lmerTest (3.1-3). Visualizations were completed using ggplot2 (3.5.1), ggridges (0.5.6), and ggeffects (2.2.1).

#### 2.5.1 Goal 1: SCFA associations with biomarker and cognitive outcomes

242 participants with amyloid status were included in Goal 1A analysis and 186 participants with tau PET status were included in Goal 1B analysis (Fig. 1). Binomial logistic regression was used to evaluate the effects of fecal SCFA level and SCFA-by-sex interactions on A+ risk (Goal 1A) and T+ risk (Goal 1B). Models were assessed separately for each pairwise combination of biomarker outcome and SCFA predictor, resulting in six models for Goal 1A and six models for Goal 1B. Outcomes were coded as binary variables to indicate positive (A+ or T+ = 1) and negative (A- or T- = 0) biomarker status. For each SCFA, participants’ levels were normalized to the cohort median, and median-normalized SCFA levels were modeled continuously. SCFA-by-sex interaction effects were tested to compare SCFA effects on males vs. females; because the analysis cohort had a greater proportion of females (Table 1), the reference contrast was defined as females with median-normalized SCFA levels of 0. Model covariates included age at fecal sample, BMI, *APOE* ε4 carrier status, clinical diagnosis, BSS, and time between biomarker and fecal sampling (i.e., age at fecal sample minus age at biomarker measurement). Age and time between biomarker and fecal sampling variables were modeled continuously and odds ratios are reported as risk per 1-year increments. *APOE* ε4 carrier status was modeled as a binary variable using ε4 noncarrier status as the reference contrast. Clinical diagnosis was modeled as a binary variable (mild cognitive impairment/dementia vs. cognitively unimpaired) using cognitively unimpaired status as the reference contrast. BSS and BMI were modeled continuously and odds ratios are reported per 1-level increments along the Bristol scale and 1 kg/m^2^ increments for BMI, respectively. Stepwise model selection was used to identify the model with the fewest terms that significantly predicted biomarker positive status. We included our predictors of interest (SCFA, sex, and SCFA-by-sex interaction) in the base model, and after each round of selection we evaluated goodness of fit with the likelihood ratio test and the Akaike information criterion (AIC). Covariates were excluded if they did not significantly decrease the log-likelihood of the model or increase its AIC. The final equation identified with model selection was biomarker (A or T) status ∼ SCFA + sex + SCFA-by-sex + age + (*APOE* ε4 carrier status) + diagnosis + (time between biomarker and fecal sampling). These models were also re-evaluated within the cognitively unimpaired subset of the analysis cohort (n=225) to assess whether SCFA levels and SCFA-by-sex interactions were significantly associated with A+ and T+ risk before the onset of cognitive decline. We reported exponentiated odds ratios and their 95% confidence interval, and we tested *P* values with Wald’s test. Effects were significant if they met a two-sided alpha of 0.05, and we controlled for Type I error by applying Benjamini-Hochberg adjustments across the six tests performed per biomarker outcome.

**Table 1.**
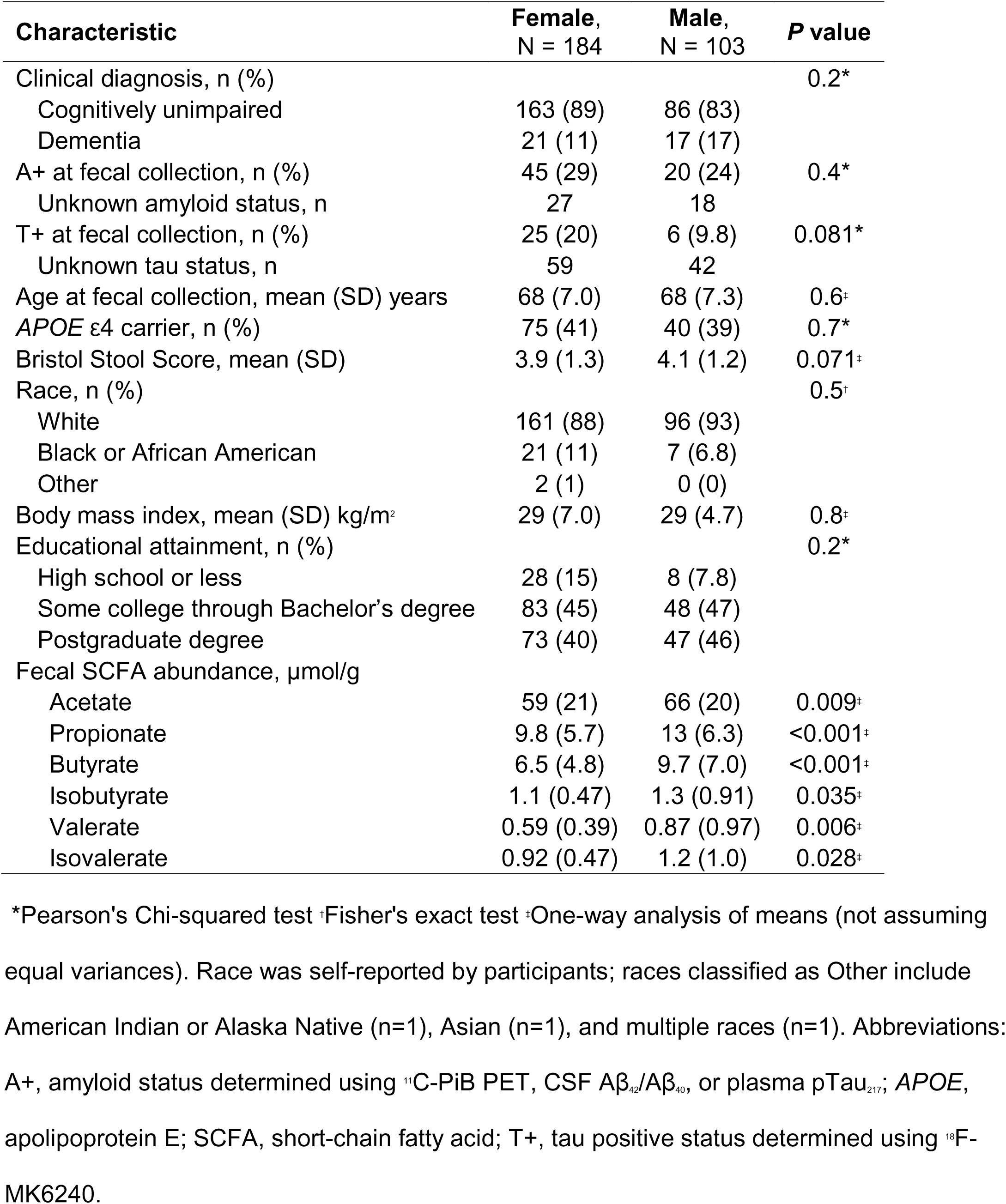
Demographics for the full participant cohort.

127 participants with CSF AD biomarkers were included in Goal 1C analyses (Fig. 1) focused on associations of SCFA levels and SCFA-by-sex interactions with CSF biomarkers. Linear regression models were tested separately per biomarker/SCFA combination, yielding a total of 54 models (9 biomarkers x 6 SCFAs). Stepwise model selection was completed as described in Goals 1A and 1B, starting with a base model of biomarker ∼ SCFA + sex + SCFA-by-sex and assessing whether covariates of age at fecal sample, BMI, *APOE* ε4 carrier status, diagnosis, BSS, time between biomarker and fecal sampling, and CSF biomarker batch improved model goodness of fit. The final equation identified in model selection was biomarker ∼ SCFA + sex + SCFA-by-male sex + age + (*APOE* ε4 carrier status) + diagnosis + (BSS) + (CSF biomarker batch). Because ordinary least squares regression resulted in heteroscedastic and non-normally distributed model residuals, we estimated parameters with maximum likelihood estimation, reporting robust 95% confidence intervals and *P* values generated with the Satorra-Bentler scaled Chi-squared test. Effects were considered significant if they met a two-sided alpha of 0.05 after FDR correction (completed as described in Goals 1A and 1B). Models were tested in the full cohort and repeated in the cognitively unimpaired subset (n=119) to determine whether effects were present before dementia onset.

Participants with longitudinal plasma pTau_217_ (n=283) and cognitive (n=219) data were included in Goals 1D and 1E (Fig. 1), which assessed whether SCFA levels at baseline predicted trajectories of biomarker accumulation or cognitive decline. Participants had plasma pTau_217_ data at 2-7 time points and PACC3 scores calculated at 2-14 time points. Linear mixed effects models were applied to assess whether SCFA or SCFA-by-sex terms modified participant pTau_217_ and cognitive slopes, running one model per outcome/SCFA pair for a total of 12 models (2 outcomes x 6 SCFAs). The model was specified as pTau_217_ or PACC3 ∼ SCFA + age + age^2^ + sex + SCFA*sex + SCFA*age + SCFA*age^2^ + sex*age + sex*age^2^ + SCFA*sex*age + SCFA*sex*age^2^ + (APOE ε4 carrier status) + BSS + (person-level random effect). If SCFA*sex*age^2^ effects were found to be nonsignificant, prespecified exploratory analyses tested effects without sex interactions in the model pTau_217_ or PACC3 ∼ SCFA + age + age^2^ + sex + SCFA*age + SCFA*age^2^ + (APOE ε4 carrier status) + BSS + (person-level random effect). Variables were defined as described in Goal 1A, and age was centered on the cohort mean baseline age and normalized on the baseline age standard deviation. Benjamini-Hochberg FDR was used to correct for inflated Type I error; effects were considered significant at an FDR-corrected alpha of 0.05, and *P* values were tested with Wald’s test. Regressions were run on the full cohort and repeated on the cognitively unimpaired subset (n=219 and 245 for pTau_217_ and cognitive analyses, respectively). Significant effects were visualized using SCFA tertiles defined within the analysis cohorts to indicate effects on outcomes at low, intermediate, and high levels of SCFAs.

#### 2.5.2 Goal 2: Microbiome associations with biomarker outcomes

287 participants were included in analyses for Goals 2A and 2B (Fig. 1) focused on MAG relationships with A+ and T+ risk; analyses were repeated in the CU subset (n=249) to determine effects in preclinical participants. We focused analyses on MAGs that encode the production of propionate and butyrate because they are the two most abundant SCFAs in the gut and are exclusively produced by gut microbes. To test the hypothesis that abundance of MAGs with propionate and butyrate production pathways is negatively associated with risk of biomarker positivity, we used binomial logistic regression as in Goals 1A and 1B, with A and T status as outcomes, and MAG levels as predictors of interest. The model from Goal 1B was used to control for clinical and sample covariates. One model was used per MAG-biomarker combination, resulting in a total of 2928 models (2 outcomes x 1464 MAGs). Benjamini-Hochberg FDR was used per biomarker outcome to correct for inflated Type I error; effects were considered significant at an FDR alpha of 0.05. Since we anticipated that the gut microbiome’s impact on AD likely stems from the cumulative function of many SCFA-producing microbes, we summarized the mean MAG effects on A+ and T+ status using Wilcoxon rank-sum tests. Tests compared odds ratios from propionate- or butyrate-producing MAGs to odds ratios from nonproducer MAGs, and effects were considered significant if they reached a two-sided alpha of 0.05. To explore whether sex differences in these MAGs could explain the observed sex-specific SCFA associations with biomarkers, we also tested Bray-Curtis dissimilarity across sex using PERMANOVA and evaluated propionate- and butyrate-producer MAG associations with sex controlling for covariates using linear regression. We applied the model MAG ∼ sex + age + BSS to each MAG separately for a total of 1464 models, and we considered associations significant if effects reached an FDR-corrected alpha of 0.05.

127 participants were included in analyses for Goal 2C focused on MAG associations with CSF biomarkers; analyses were also repeated in the CU subset (n=119). To test the hypothesis that abundance of MAGs with propionate and butyrate production pathways is negatively associated with CSF biomarker levels, we applied multiple linear regression that included CSF biomarkers as outcomes, and SCFAs and SCFA-by-sex as predictors of interest. The model from Goal 1C was used to control for appropriate covariates. One model was used per outcome, for a total of 13176 models (9 CSF biomarkers x 1464 MAGs). Regressions were performed as done for Goal 1C, using maximum likelihood estimation and Satorra–Bentler corrections to adjust for heterogeneity of model residuals. Benjamini-Hochberg FDR was used to correct for inflated Type I error across groups of 1464 models that shared a biomarker outcome (FDR = 0.05). Mean MAG effects were summarized using Wilcoxon rank-sum tests as described above.

#### 2.5.3 Goal 3: SCFA mediation of relationships between the gut microbiome and biomarkers

249 CU participants were included in analyses for Goal 3. We applied path analysis with nonparametric bootstrapping (1,000 simulations) using the mediate() function from the mediation R package. Models were evaluated for all MAGs regardless of whether they had genes for SCFA production, because some microbes support SCFA production indirectly through microbiome cross-feeding (e.g., producing a SCFA precursor molecule which is later metabolized by a direct SCFA producer). Models were evaluated per MAG to identify the potential impact of specific bacteria in the microbiome. We defined MAG abundance as the treatment variable, SCFA level as the mediator, and amyloid status or CSF biomarkers with significant relationships to SCFAs as the outcome variables. Covariates for both the direct effect and mediation effect models included age, sex, *APOE* ε4 carrier status, time between fecal sample and biomarker outcome, and BSS. The proportions of mediation effects were calculated by average causal mediation effects (ACME)/ average causal mediation effects (ACME) + average direct effects (ADE). For the MAG-biomarker relationships with significant SCFA mediation effects (proportion of mediated effects *p*<.05), we reported whether the MAGs contained genes encoding propionate or butyrate production, and we characterized the direction of the indirect relationships (i.e., MAG-SCFA and SCFA-biomarker).

## 3. Results

### 3.1. Participant sample characteristics

Table 1 describes the demographics for the full cohort. Among the 287 participants, 163 females and 86 males were CU, and 21 females and 17 males had dementia. 65 participants (53 CU) were A+ at the time of fecal collection; amyloid status was determined using PET, CSF, and plasma in 141, 60, and 86 participants, respectively. No clinical or demographic characteristics differed significantly by sex, however all six SCFAs were present at higher levels in males compared to females (Supplementary Fig. 3; *P* value range: <.001-.035). This and prior literature suggesting sex-specific effects of the gut microbiome on health outcomes[22] motivated tests of SCFA-by-sex interactions on AD pathology and cognitive outcomes. Across the full cohort, mean fecal SCFA levels (Table 1) were close to the range of expected physiological levels, as previous studies have reported human fecal SCFA levels of ∼60 g/kg for acetate, 10–20 g/kg for propionate, and 3.5–32.6 g/kg for butyrate (approximately 60:20:20 ratio) in healthy, middle-aged participants[43]. Compared to the full cohort (Supplementary Table 3) and the subset with longitudinal cognitive data (Supplementary Table 4), the participant sets with biomarker data had a smaller proportion of individuals with dementia (*p*=.012 and .002, respectively), but had no other significant differences in other demographics.

Supplementary Table 1 compares the medication usage between CU A-, CU A+, and AD participants. Of these medications, anticholinesterases, anticonvulsants, selective serotonin reuptake inhibitors (SSRIs), atypical antipsychotics, memantine, statins, and vitamin E differed between groups. AD medications (anticholinesterases, memantine) were exclusively found among AD participants, and a small proportion of participants took anticonvulsants, atypical antipsychotics, or vitamin E. A higher percentage of AD participants were taking statins or SSRIs, which is consistent with the high prevalence of cardiovascular disease and depression in individuals with AD. Given that observed differences in medication usage largely reflected underlying clinical diagnoses and no unexpected or disproportionate medication patterns were identified, medications were not included as covariates in analyses.

### 3.2. Fecal levels of propionate and isovalerate are associated with lower A+ risk

We first tested for relationships between SCFAs and A+ risk. Stepwise model selection showed that the best fitting logistic regression model across both biomarker outcomes (A and T positivity status) and all SCFAs included SCFA main effects and SCFA-by-sex effects, as well as the covariates age, sex, *APOE* ε4 carrier status, diagnosis, and time between fecal sample and biomarker measurement (Supplementary Table 5). Within the full cohort, propionate and isovalerate were associated with lower A+ risk before FDR correction (Fig. 2) (propionate: odds ratio[OR] (95% CI) = 0.33 (0.12, 0.78), *p*=.02, *q*=.09; isovalerate: OR (95% CI) = 0.396 (0.16, 0.88), *p*=.03, *q*=.09). Results were consistent when restricting analyses to CU participants (Supplementary Table 6). SCFA-by-sex interaction effects were not significant in any of the twelve models for either the full cohort or the CU subset. SCFA and SCFA-by-sex odds ratios were consistent in their magnitude, direction and significance in sensitivity analyses that excluded extreme outliers (values above quartile 3 plus three times an SCFA’s interquartile range; n=0-4 per SCFA). We observed no significant associations between SCFAs and T+ risk (Supplementary Fig. 4, Supplementary Tables 7-8).

**Figure 2.**
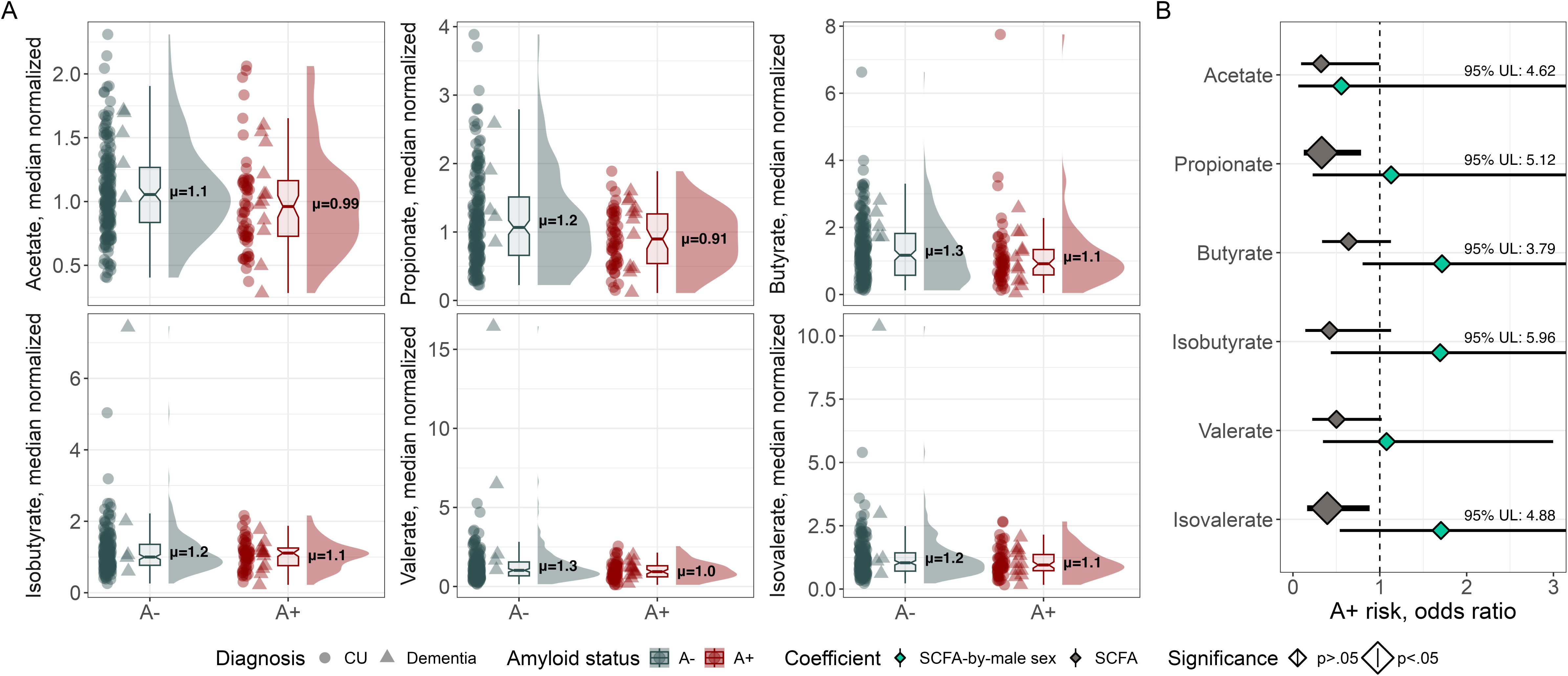
Amyloid relationships with SCFA abundance. (**A**) Fecal SCFA levels compared across A+ and A-participants and (**B**) forest plot with odds ratios depicting A+ risk as a function of SCFA level and modified by male sex. From left to right per panel, a scatter plot depicts individual participant values, a box plot shows median SCFA level per amyloid status, and a density plot shows the data distribution and highlights the mean SCFA level per amyloid status. SCFA levels in CU participants are depicted with circles, and in MCI or dementia participants with triangles. For each SCFA, abundances are normalized on the median abundance represented in the sample. Odds ratios for main SCFA effects and SCFA-by-sex interactions were derived from the logistic regression model *Amyloid status ∼ SCFA + sex + SCFA-by-sex + age + APOE* ε*4 carrier status + diagnosis + time between biomarker measurement and fecal collection*. To allow for visual comparison of mean effects, the 95% confidence interval upper limits were annotated for coefficients with wide confidence intervals. Abbreviations: A+/-, amyloid positive or negative status determined with ^11^C-PiB PET, CSF Aβ_42_/Aβ_40_, or plasma pTau_217_; *APOE*, apolipoprotein E; CU, cognitively unimpaired; MCI, mild cognitive impairment; SCFA, short-chain fatty acids; UL, upper limit of the 95% confidence interval.

### 3.3. SCFA and SCFA-by-sex are associated with CSF biomarkers of amyloid and microglial activation

With CSF data, we tested the hypothesis that SCFAs are inversely associated with CSF biomarkers of AD, neuroinflammation, and neurodegeneration in a sex-specific manner (Supplementary Table 9). Among nine biomarkers, Aβ_42_/Aβ_40_ and sTREM2 were significantly associated with SCFA levels after FDR correction (Fig. 3A). The β coefficient represents SCFA effects on CSF biomarkers. In CU participants, isobutyrate and isovalerate correlated inversely with Aβ_42_/Aβ_40_ in males but not females (isobutyrate-by-male sex β (95% CI) = -0.016 (-0.03, - .005), *q*=.014; isovalerate-by-male sex β (95% CI) = -0.014 (-0.02, -.004), *q*=.014) (Fig. 3B-C). Butyrate was also correlated with lower sTREM2 in males compared to females (butyrate-by-male sex β (95% CI) = -0.004 (-1.7, -0.3), *q*=.032) (Fig. 3D). Higher acetate, butyrate, isobutyrate, and isovalerate levels showed significant main effects on higher Aβ_42_/Aβ_40_ (acetate β (95% CI) = 0.011, *q*=.043; butyrate β (95% CI) = 0.005 (9.28e-4, 0.0096), *q*=.043; isobutyrate β (95% CI) = 0.008 (7.92e-4, 0.01), *q*=.043; isovalerate β (95% CI) = 0.007 (8.23e-4, 0.01), *q*=.043) (Fig. 3E-H). No significant SCFA effects were observed on GFAP, neurogranin, NfL, pTau_181_/Aβ_42_, tTau, YKL-40, or ⍺-synuclein (Fig. 3). Results in the CU subset reflect those in the full cohort (Supplementary Fig. 5, Supplementary Table 10).

**Figure 3.**
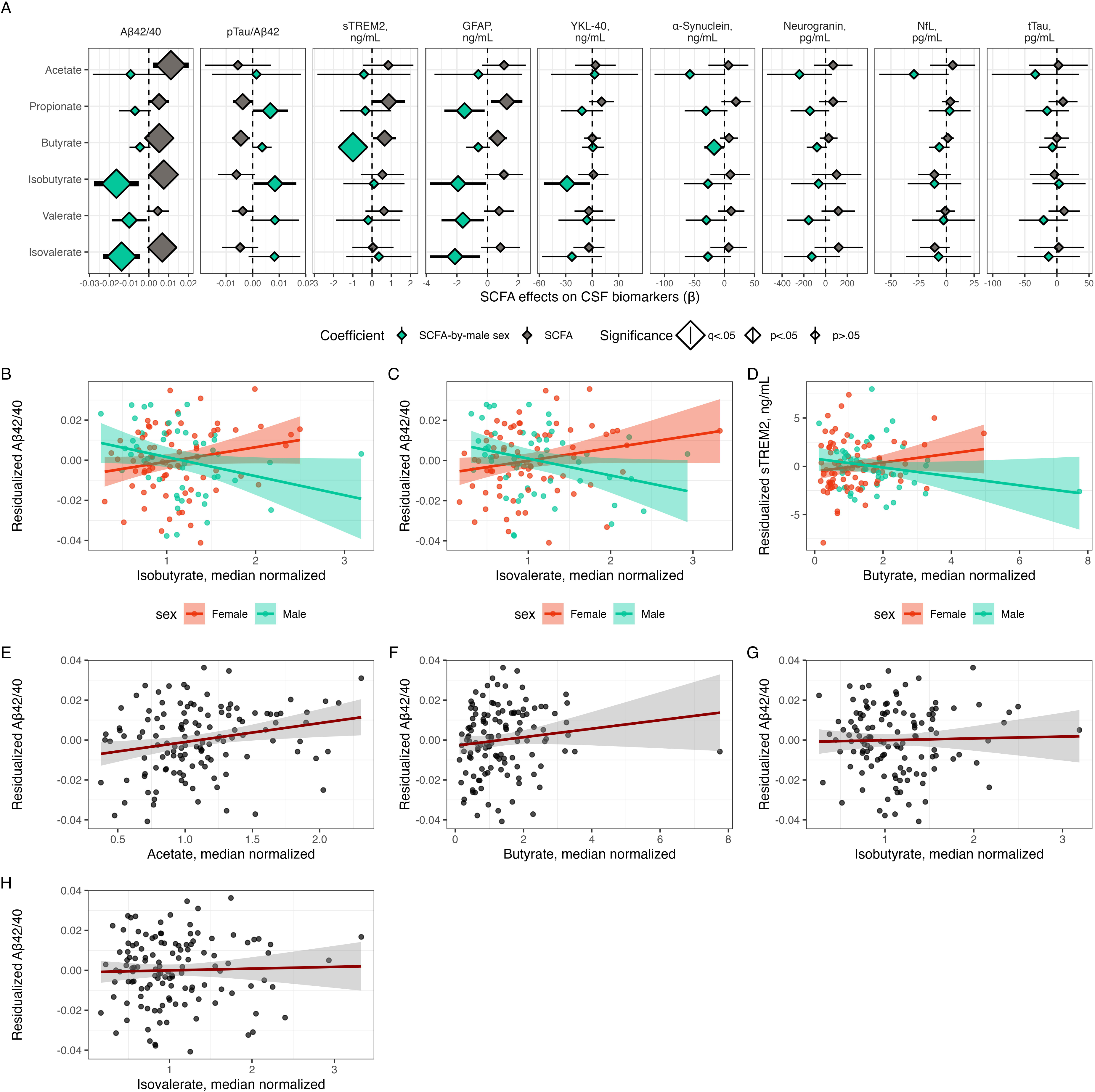
SCFAs significantly associated with CSF biomarkers of amyloid pathology and microglial activation in cognitively unimpaired participant subset. (**A**) Forest plot with SCFA and SCFA-by-male sex effect coefficients on CSF biomarker outcomes. Effects were estimated with the regression model *CSF biomarker ∼ SCFA + sex + SCFA-by-sex + age + APOE* ε*4 carrier status + BSS + CSF biomarker batch*. (**B-D**) CSF biomarker levels residualized on model covariates (age, sex, SCFA level, *APOE* ε4 carrier status, BSS, CSF biomarker batch) to demonstrate (**B-C**) Aβ_42_/Aβ_40_ relationships with isobutyrate and isovalerate and (**D**) sTREM2 relationships with butyrate. (**E-H**) Aβ_42_/Aβ_40_ residualized on model covariates (age, sex, *APOE* ε4 carrier status, BSS, CSF biomarker batch) to demonstrate amyloid relationships with (**E**) acetate, (**F**) butyrate, (**G**) isobutyrate, and (**H**) isovalerate main effects. Abbreviations: Aβ, amyloid-beta; *APOE*, apolipoprotein E; BSS, Bristol Stool Score; GFAP, glial fibrillary acidic protein; NfL, neurofilament light chain protein; pTau_181_, tau phosphorylated at amino acid 181; SCFA, short-chain fatty acids; sTREM2, soluble triggering factor expressed on myeloid cells 2; tTau, total tau; YKL-40, chitinase-3-like protein 1.

### 3.4. Higher SCFA levels are associated with slower plasma pTau_217_ accumulation in females and slower cognitive decline

In addition to cross-sectional SCFA relationships with pathology, we tested whether SCFA levels at baseline were associated with slopes of plasma pTau_217_ accumulation and decline on PACC3 performance. In the CU subset we observed significant sex-specific effects of valerate (Fig. 4A) and isovalerate (Fig. 4B) on plasma pTau_217_ trajectories (Supplementary Table 11), such that higher SCFA levels were associated with slower pTau_217_ accumulation in female participants but not males (valerate SCFA*male sex*age^2^ β (95% CI) = 0.04 (2.2e-4, 0.07), *p*=.049, *q*=.15; isovalerate β (95% CI) = 0.06 (0.02, 0.099), *q*=.02). These results were also reflected in the full cohort (Supplementary Fig. 6A-6B, Supplementary Table 12). SCFAs had no sex-specific effects on PACC3 trajectories in the CU cohort, however all six SCFAs had significant main effects (Fig. 4C-H) such that participants with lower SCFA levels had faster cognitive decline (SCFA*age^2^ *q*s = .0004-.0024; Supplementary Table 13). Lower valerate and isovalerate in the full cohort were significantly associated with faster PACC3 decline (Supplementary Fig. 6C-6D; valerate SCFA*male sex*age^2^ β (95% CI) = -0.1 (-0.2, -0.008), *p*=0.03, *q*=.1; isovalerate β (95% CI) = -0.15 (-0.27, -0.02), *p*=.02, *q*=.1), although results did not survive FDR correction (Supplementary Table 14).

**Figure 4.**
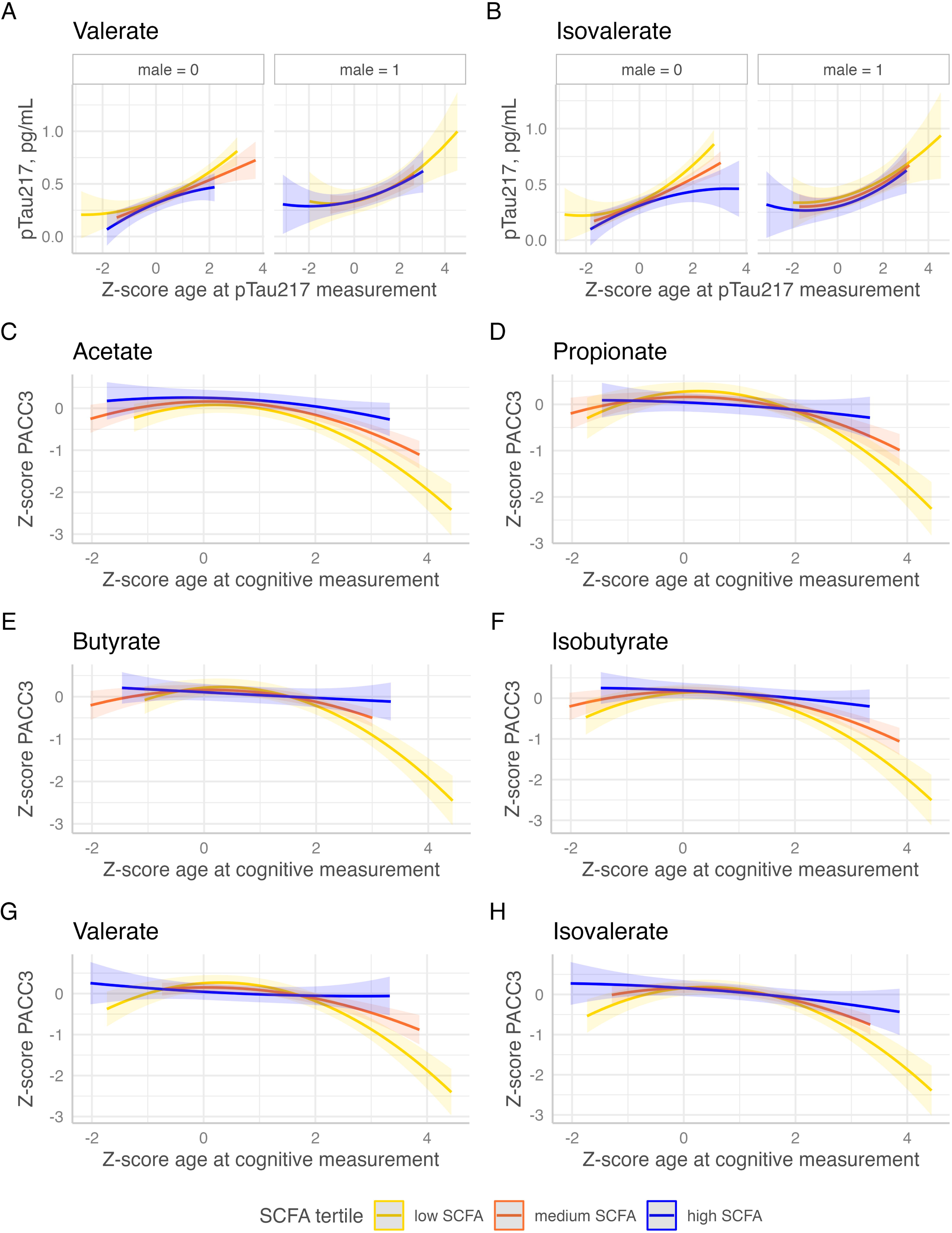
Effects of SCFAs on longitudinal trajectories of pTau_217_ and cognition in cognitively unimpaired subset. (**A**) Effects of SCFAs on longitudinal trajectories of plasma pTau_217_ or (**B**) cognition (PACC3) by multiple linear regression: *plasma pTau_217_ or cognition ∼ SCFA + sex + SCFA-by-male sex + age^2^ + APOE* ε*4 carrier status + BSS*. For visualization, SCFA levels (normally distributed) are grouped into even tertiles: high, medium, and low. Abbreviations: *APOE*, apolipoprotein E; BSS, Bristol Stool Score; PACC3, Preclinical Alzheimer Cognitive Composite score (three item version); SCFA, short-chain fatty acids.

### 3.5. MAGs harboring key genes for SCFA production are associated with lower A+ risk

To investigate relationships between biomarkers and gut microbes with genes for production of propionate or butyrate, we tested associations between microbial genes and A or T status. Reference-based mapping of sequences to MetaCyc pathways showed that a propionate production pathway was one of the pathways most associated with lower risk of being A+ (OR (95% CI) = 0.52 (0.32, 0.85), *p*=.01, *q*=.21; Supplementary Fig. 7-8, Supplementary Tables 15-16). However, previous work shows that this method is not ideal for quantifying relative abundance of propionate and butyrate production pathways[45], so *de novo* metagenome assembly was used to generate MAGs. Since *de novo* assembly is largely unconfirmed with annotation, we tested whether MAG levels represented SCFA levels in the same sample; as anticipated, we observed that MAGs with butyrate and propionate production genes had higher relative abundances in samples with high levels of butyrate and propionate, respectively (Supplementary Fig. 9). MAGs with genes for butyrate production primarily included bacteria in the families *Lachnospiraceae* and *Ruminococcaceae*, both within the phylum Bacillota (previously known as Firmicutes) (Supplementary Table 17). MAGs with genes for propionate production primarily included microbes in the phyla Bacteroidota (previously known as Bacteroidetes) and Bacillota, particularly *Bacteroidaceae* and *Lachnospiraceae* (Supplementary Table 17).

Although PERMANOVA testing Bray-Curtis dissimilarity did not reveal broad compositional differences in MAGs between A+ and A-participants (Supplementary Fig. 10), Wilcoxon rank-sum testing showed that MAGs with propionate production genes were significantly associated (*p*=1.1e-5) with lower risk of A+ status (log transformed odds ratio (μ)=-0.045, mean OR=0.96) compared to MAGs without these genes (μ=0.004, mean OR=1.004) (Fig. 5A, Supplementary Table 17). Similarly, MAGs with butyrate production genes showed a nonsignificant (*p*=.11) association with lower A+ risk (μ=-0.024 vs. -0.002 and mean OR=0.97 vs. 0.99 for MAGs with and without butyrate production genes, respectively) (Fig. 5B). Similar results were observed in the full cohort (Supplementary Fig. 11, Supplementary Table 18). Additionally, although we did not observe broad compositional differences in MAGs between T+ and T-participants (Supplementary Fig. 12), we found that MAGs with genes for propionate or butyrate production were associated with lower T+ risk compared to MAGs without those genes (Supplementary Fig. 13; *p*=9.5e6 and .00015 when comparing propionate- and butyrate-producing MAGs, respectively, to nonproducers). Analysis of the overall microbiome composition also showed significant sex differences in MAG abundance (*p*=.014) (Supplementary Fig. 14). MAGs with propionate-producing genes showed no significant differences in their abundance between males and females (*p*=.074, Supplementary Fig. 15A, C), however butyrate-producing MAGs were more abundant in males compared to females, in contrast with butyrate nonproducers which showed almost no sex differences in abundance (*p*<2.22e-16, Supplementary Fig. 15B, D).

**Figure 5.**
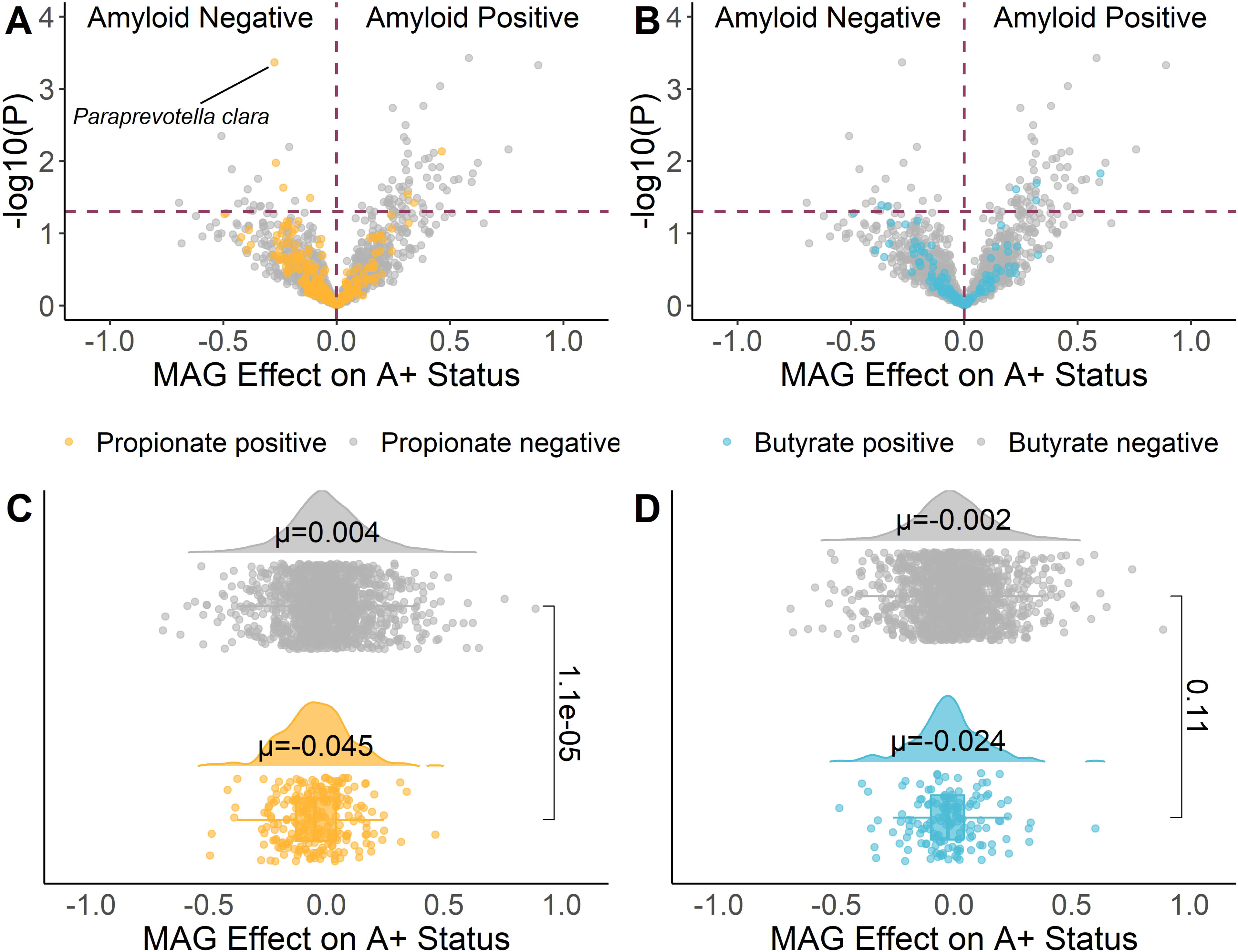
Distributions of propionate- and butyrate-producing bacteria among cognitively unimpaired participants are shifted toward amyloid-negative status. The distribution of log transformed odds ratios (μ) for metagenome-assembled genomes (MAGs) with genetic pathways for (**A**,**C**) propionate production and (**B**,**D**) butyrate production in A+ participants compared to A-, for producer MAGs vs. non-producer MAGs (labeled propionate or butyrate positive or negative by color), as assessed by a Wilcoxon rank-sum test for log transformed odds ratios, indicating the effect of each MAG on amyloid status (negative values = A-, positive values = A+). Effects of MAGs on amyloid status were assessed by logistic regression: *Amyloid status ∼ MAG + sex + age + APOE* ε*4 carrier status + BSS.* (**C**,**D**): From top to bottom per panel, a density plot shows the data distribution, highlighting the mean MAG log transformed odds ratio per group, and a scatter plot depicts the log transformed odds ratio for each MAG. Propionate positive and Butyrate positive MAGs were classified based on gene content. Abbreviations: A+/-, amyloid positive or negative status determined with ^11^C-PiB PET, CSF Aβ_42_/Aβ_40_, or plasma pTau_217_; *APOE*, apolipoprotein E; BSS, Bristol Stool Score; MAG, metagenome-assembled genome.

### 3.6. MAGs with genes for propionate and butyrate production are associated with CSF biomarkers of AD pathology, neuroinflammation, and neurodegeneration

We tested whether MAGs with genes for propionate or butyrate production have associations with CSF biomarkers, using linear regressions models that included MAG-by-sex interactions and controlled for the same covariates as in Goal 1B (Supplementary Table 19). Since we observed that SCFAs have associations with Aβ_42_/Aβ_40_ and sTREM2, here we highlight associations of propionate- and butyrate-producing MAGs with Aβ_42_/Aβ_40_ and sTREM2. Higher abundances of propionate-producing MAGs were associated with low Aβ_42_/Aβ_40_ in females (mean MAG β=-0.005, *p*=.0024), and high Aβ_42_/Aβ_40_ in males (mean MAG-by-male sex β=0.019, *p*=.0056) (Fig. 6A). In contrast, high butyrate-producing MAG abundance was associated with high Aβ_42_/Aβ_40_ in females (mean MAG β=0.022, *p*<2.22e-16), with an attenuated effect in males (mean MAG-by-male sex β=-0.003, *p*=.00065) (Fig. 6B).

**Figure 6.**
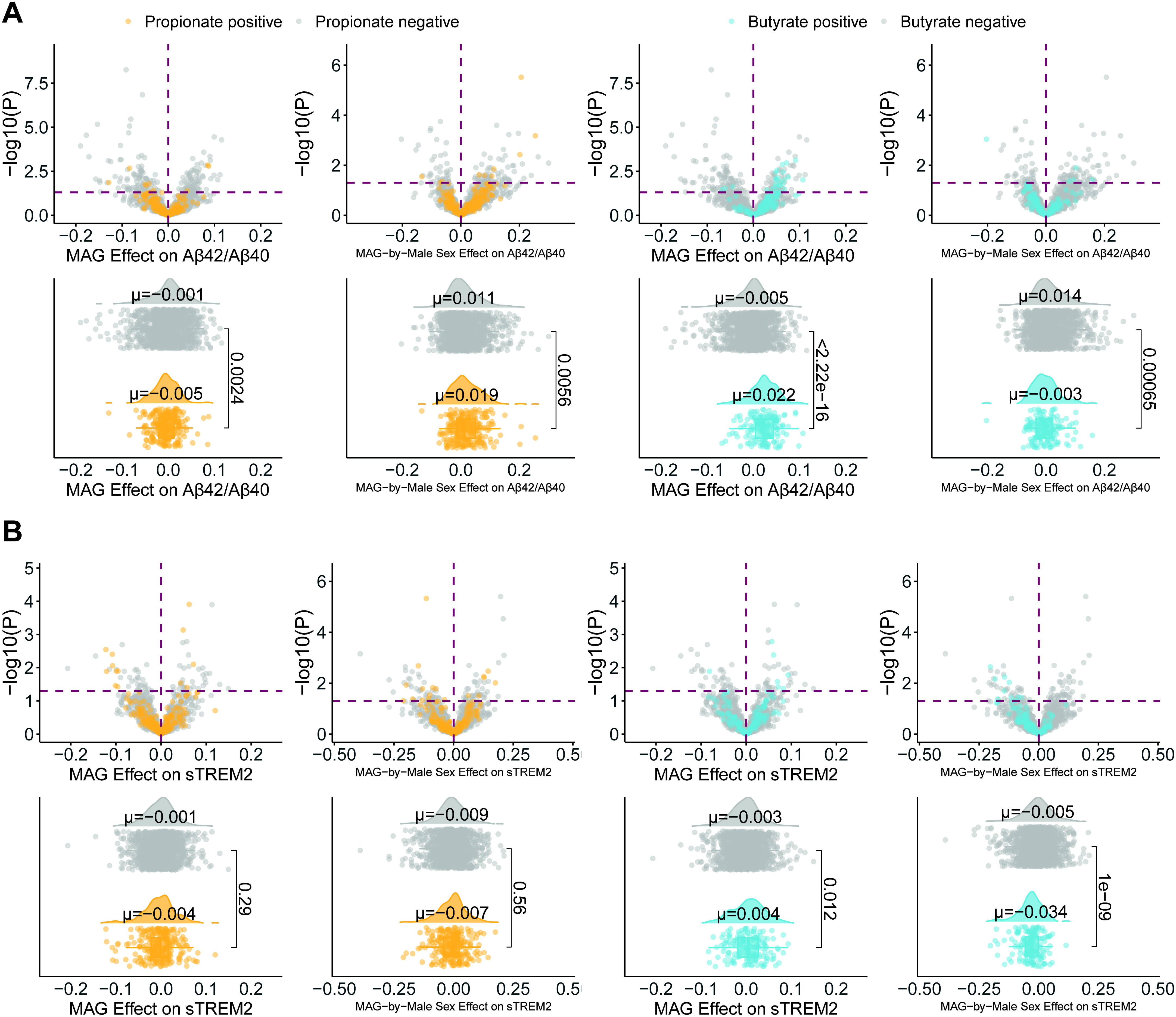
MAGs significantly associated with CSF biomarkers of AD, neuroinflammation, and neurodegeneration in cognitively unimpaired participant subset. Propionate- and butyrate-producing MAGs, as well as MAG-by-male sex interaction terms, have effects on CSF (**A**) Aβ_42_/Aβ_40_ and (**B**) sTREM2, as assessed by multiple linear regression: *CSF biomarker ∼ MAG + sex + MAG-by-male sex + age + APOE* ε*4 carrier status + BSS + CSF biomarker batch*. *P* values shown are before FDR correction. Abbreviations: Aβ_42_/Aβ_40_, amyloid beta 42 to amyloid beta 40 ratio; *APOE*, apolipoprotein E; BSS, Bristol Stool Score; GFAP, glial fibrillary acidic protein; MAG, metagenome-assembled genome; NfL, neurofilament light chain protein; pTau_181_/Aβ_42_, phosphorylated tau 181 to amyloid beta 42 ratio; sTREM2, soluble triggering factor expressed on myeloid cells 2; tTau, total tau; YKL-40, chitinase-3-like protein 1; SCFA, short-chain fatty acid.

In addition to effects with amyloid, we found that higher butyrate-producing MAG abundances were associated with higher sTREM2 in females (mean MAG β=0.004, p=.012) and with lower sTREM2 in males (mean MAG-by-male sex β=-0.034, p=1e-9) (Fig. 6D). Propionate-producing MAGs did not show significant associations with sTREM2 (Fig. 6C). Beyond Aβ_42_/Aβ_40_ and sTREM2, butyrate- and propionate-producing MAGs showed significant main effects with higher pTau181/Aβ42, GFAP, neurogranin, NfL, and YKL-40; propionate producers had significant effects with tTau and α-synuclein, however butyrate producers did not. MAG-by-sex effects also showed associations with lower biomarker levels in males, although sex-specific associations of butyrate producers with tTau and of propionate producers with YKL-40 and α-synuclein were not significant. Results for the CU cohort (Supplementary Table 20) were similar to those in the full cohort (Supplementary Tables 21-22).

### 3.7. Fecal propionate mediates the relationship between propionate-producing bacteria and AD biomarkers

Because propionate, isovalerate, and propionate-producing microbes were significantly associated with lower A+ risk, we tested whether fecal propionate and isovalerate levels mediated associations between MAGs and A+ status (Fig. 7A-B). Propionate significantly mediated 58 MAG relationships with A+ risk (FDR *p*<.05); 11 of the MAGs involved had genes that encode for propionate or butyrate production, and 45 MAGs had positive associations with A+ status (Fig. 7A). Fig. 7C illustrates the mediation effect that propionate showed between *Paraprevotella clara* and A+ risk (Supplementary Table 23). *P. clara* was negatively associated with A+ risk (direct effect β=-0.023, *p*<.001), and propionate was a significant mediator of the total effect (proportion mediated=13.5%). Propionate also significantly mediated the negative associations of twelve other MAGs – *Dialister sp000434475,* two strains of *Roseburia inulinivorans, Dorea formicigenerans, Enterocloster sp000431375, Merdisoma sp 900553635, Phascolarctobacterium A sp900544885, Coprobacter secundus, Emergencia, UBA1394 sp900554975,* and two strains of *Copromonas sp900066535* – with A+ risk (Fig. 7A, Supplementary Table 23). Because we also observed that isovalerate was significantly associated with lower A+ risk, we additionally tested whether fecal isovalerate levels mediated associations between MAGs and A+ status. *Ruminococcus D bicirculans* was negatively associated with A+ risk (direct effect β=-0.032, *p*=.004), and isovalerate mediated -0.169% of the total effect through a positive association with *Ruminococcus D bicirculans* and a negative association with A+ risk (Fig. 7A, Supplementary Table 23).

**Figure 7.**
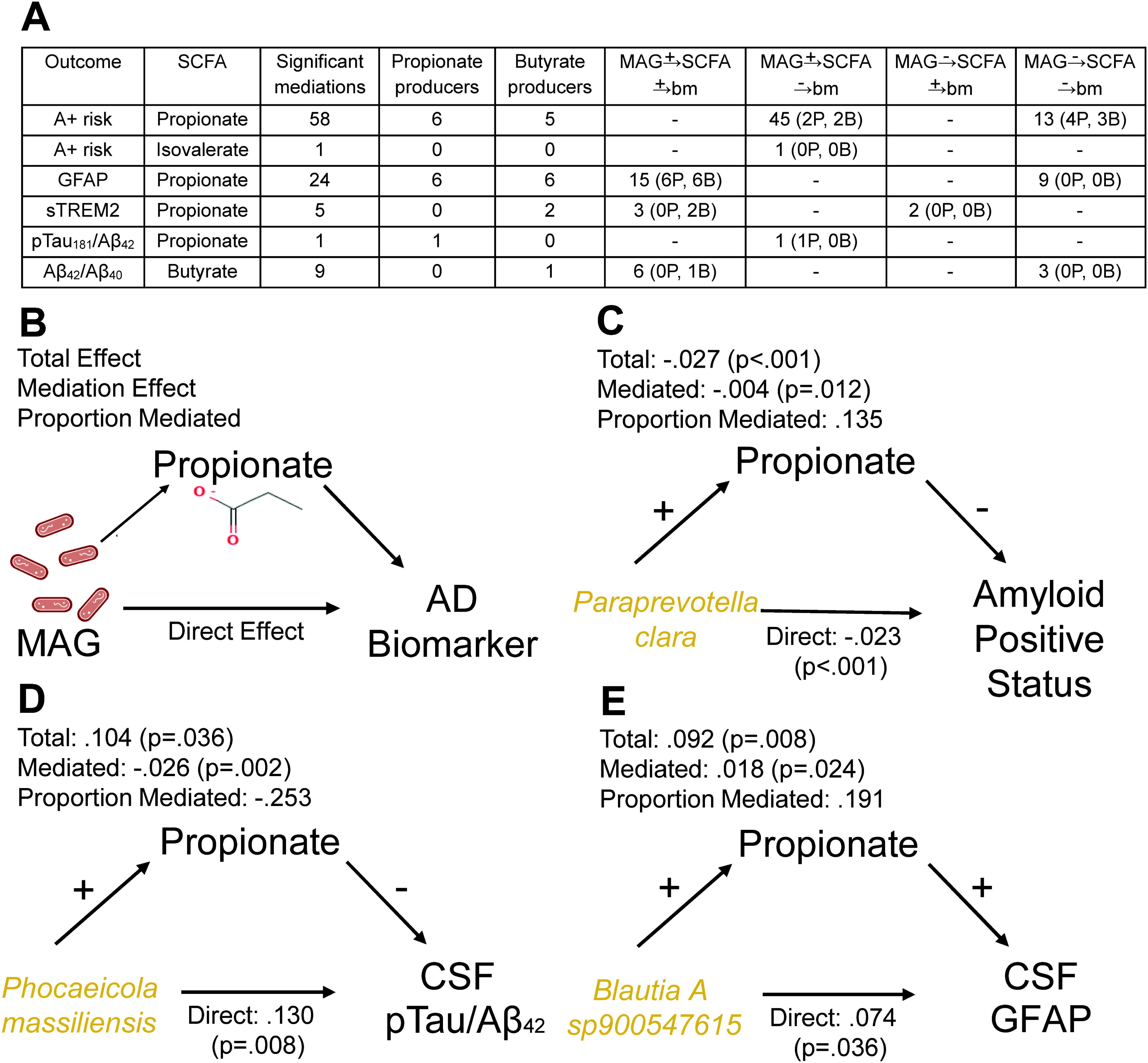
SCFA mediation of relationship between MAGs and AD biomarkers. (**A**) Table of significant mediation relationships. Mediation assessed by (**B**) causal mediation analysis, accounting for the covariates sex, age, APOE ε4 carrier status, BSS, and CSF biomarker batch. (**C**) Propionate mediates relationship between *Paraprevotella clara* and A+ status. Propionate mediates relationship between propionate-producing MAGs and (**D**) CSF pTau_181_/Aβ_42_, and (**E**) CSF GFAP. Effects of SCFAs on CSF biomarkers were controlled for sex, age, *APOE* ε4 status, and BSS. Propionate producing and Butyrate producing MAGs were classified based on gene content. Yellow = propionate-producing MAG. Abbreviations: A+, amyloid positive status determined with ^11^C-PiB PET, CSF Aβ_42_/Aβ_40_, or plasma pTau_217_; Aβ_42_/Aβ_40_, amyloid beta 42 to amyloid beta 40 ratio; *APOE*, apolipoprotein E; B, butyrate producer; bm, biomarker; BSS, Bristol Stool Score; GFAP, glial fibrillary acidic protein; MAG, metagenome-assembled genome; P, propionate producer; pTau_181_/Aβ_42_, phosphorylated tau 181 to amyloid beta 42 ratio; SCFA, short-chain fatty acid; sTREM2, soluble triggering factor expressed on myeloid cells 2.

We applied the same path analysis models to test SCFAs as mediators of relationships between MAGs and CSF biomarkers of pathology (Fig. 3). Propionate was a significant mediator of 24 MAG relationships with GFAP, 11 of which produced propionate or butyrate (one MAG harbored genes encoding production of both SCFAs) (Fig. 7A). *Phocaeicola massiliensis* was positively associated with pTau_181_/Aβ_42_ (direct effect β=0.130, *p*=.008), and propionate mediated -25.3% of the total effect through a positive association with *P. massiliensis* and a negative association with pTau_181_/Aβ_42_ (Fig. 7D, Supplementary Table 24). *Blautia A sp900547615* was positively associated with GFAP (direct effect β=0.074, *p*=.036), and propionate was positively associated with *Blautia A sp900547615* and positively associated with GFAP levels, mediating 19.1% of the total effect (Fig. 7E, Supplementary Table 24). Propionate also showed mediation effects for the positive relationships of 14 other MAGs with GFAP (Three strains of *Faecalibacterium prausnitzii, Holdemanella biformis,* two strains of *Roseburia inulinivorans, Blautia hansenii, Brachyspira aalborgi, UBA1394 sp900554975,* a strain of *UBA7173*, *Roseburia sp900753715,* a strain of *CAG-873,* a strain of *CAG-485, Ruminococcus B gnavus*) (Fig. 7A, Supplementary Table 24). Propionate also mediated associations of five MAGs (two butyrate producers) with sTREM2, and of one MAG (a propionate producer) with pTau_181_/Aβ_42_(Fig. 7A, Supplementary Table 24). Butyrate mediated the positive association between six MAGs (one butyrate producer) and Aβ_42_/Aβ_40_ (*Faecalibacterium longum*, five strains of *Gemmiger formicilis*) (Fig. 7A, Supplementary Table 24). No mediation effects of isobutyrate or isovalerate on MAG relationships with Aβ_42_/Aβ_40_ were observed.

## 4. Discussion

In this work, we used fecal SCFAs and metagenomic deep sequencing to assess relationships of gut microbiome SCFA production with AD pathology and cognitive decline. The observed reductions in fecal propionate and isovalerate, along with propionate-producing bacteria, in A+ participants suggest a disruption in microbial metabolic function early in pathogenesis. Mediation analysis implicated propionate as a potential mechanistic link between gut microbes and AD pathology. There were also several observed sex-specific associations of SCFAs with the biomarkers Aβ_42_/Aβ_40_ and sTREM2 in CSF, and with pTau_217_ in plasma. These results support the role of microbiome-related sex differences in AD development. Further, higher SCFA levels were associated with slower cognitive decline, suggesting a functional impact of these microbial metabolites on AD progression. These findings underscore the potential of gut-mediated SCFAs as modifiable therapeutic targets.

### 4.1. Links of propionate, isovalerate, and related microbes with A+ risk

In the asymptomatic cohort, we found that propionate and isovalerate levels were lower in A+ individuals. With prior literature on SCFA roles in human health, these results suggest that SCFA changes in the gut could contribute to disease progression. Propionate is known to promote anti-inflammatory mechanisms[47–49] and ameliorate insulin resistance[43,47,50,51]. As insulin has mechanistic ties to amyloid deposition[52,53], our observations here could suggest propionate mitigates risk for amyloid pathology through modulating insulin-related pathways. However, findings are conflicted regarding the relationship between propionate and host health; high levels of propionate have been observed in AD patients’ saliva[54] and in AD mouse models’ feces and brains[55–57]. These seemingly contradictory findings may reflect differences between propionate production in the gut and its absorption and systemic bioavailability. Further work is needed to clarify the mechanisms of propionate at different sites within the host.

Because the branched-chain fatty acid (BCFA) isovalerate is less abundant in the gut environment, less is known about the dominant microbial pathways involved in its production or its impact on health outcomes. One study suggests the gene *porA* may contribute to production of the BCFAs isobutyrate and isovalerate[58]. However, there are likely other undiscovered genes that encode for BCFA production; as a result, we did not annotate or analyze pathology relationships to bacteria with these genes. Furthermore, findings are conflicted regarding BCFAs’ potential roles in AD; one study found reduced levels of these compounds in fecal samples of AD participants, while another found that fecal isovalerate and isobutyrate levels were increased in participants with dementia[12,59,60]. With previous literature suggesting propionate and isovalerate have complex relationships with human health, our current study supports their potentially beneficial effect on AD pathology and motivates future investigation into microbial production of BCFAs.

Analyses of SCFA-producing bacteria also suggested they may protect against amyloid pathology development. We observed that *Paraprevotella clara,* a Gram-negative anaerobic bacterium in the Bacteroidota phylum[61], had genes for a propionate production pathway and showed the greatest abundance reductions among A+ participants. Additionally, propionate mediated 13.5% of its relationship with amyloid status. These results contrast with previous findings that *P. clara* predicted greater clinical severity of dementia among AD participants[62]. While *P. clara* may have genes with detrimental functions at later stages of AD, our study in a predominantly cognitively unimpaired cohort suggests this bacterium and its metabolite propionate could hold promise as an intervention to potentially delay onset of amyloid pathology.

### 4.2. Sex-specific butyrate, acetate, and bacterial associations with amyloid and microglial activation biomarkers in CSF

Beyond associations with amyloid positivity, we observed relationships of SCFAs with multiple CSF biomarkers even before cognitive decline. Higher butyrate and acetate levels were both associated with lower amyloid pathology, and butyrate was linked to lower pTau_181_/Aβ_42_, indicating that these SCFAs may be linked to more favorable AD biomarker profiles. Butyrate and butyrate-producing bacteria also showed a sex-specific relationship with sTREM2, a marker of microglial activation. In females, higher butyrate levels were associated with increased activation, whereas in males, higher levels corresponded to lower sTREM2, which may reflect reduced microglial activation or a decreased number of microglia. The AD pathological implications of lower microglial activation are debated, however in preclinical AD microglia are thought to protect against amyloid aggregation[63]. Isovalerate and isobutyrate also showed sex-specific effects, wherein higher levels were associated with lower amyloid pathology in females, but higher amyloid pathology in males. Propionate- and butyrate-producing bacteria showed sex-specific relationships with CSF Aβ_42_/Aβ_40_, suggesting that propionate-producing MAGs may have a protective effect against amyloid pathology in males, and butyrate-producing MAGs may do so in females. Taken together, these results suggest that SCFAs and their bacterial producers could promote microglial response and ameliorate amyloid deposition in females, but not in males, during preclinical AD. Mediation analysis further indicates that propionate may play a mediating role in these relationships.

Relationships between sex differences in AD pathology, SCFAs, and their microbial producers may stem from hormonal factors. Several studies provide evidence that sex hormones could indirectly affect AD via effects on gut microbes. For instance, gut microbiome diversity and composition are altered with hormonal shifts across female reproductive stages; one study found that post-menopausal women have reduced levels of Bacillota (previously known as Firmicutes) including the genus *Ruminococcus*, known to include SCFA producers[64]. Other studies find that estrogen is positively associated with the abundance of SCFA-producing bacteria such as *Butyricimonas*[65], and with SCFAs themselves, as ovariectomized estrogen-deficient mice have reduced fecal butyrate[66]. Further investigation will clarify how SCFA-related mechanisms may modulate amyloid deposition and neuroinflammatory response in a sex-specific manner.

### 4.3. SCFA relationships with pTau_217_ and cognitive trajectories

In addition to cross-sectional effects with AD pathology, we found higher valerate and isovalerate levels were associated with slower pTau217 accumulation, particularly in females, and all investigated SCFAs were linked to slower cognitive decline. Because plasma pTau_217_ aggregates at a linear rate once levels have reached the positivity threshold[1], we hypothesize the slope associations indicate that higher fecal SCFA levels may decrease the likelihood of an individual becoming a pTau_217_ accumulator. The evolving literature is conflicted on whether sex differences in AD are attributable to differences in amyloid deposition; across both our cross-sectional and longitudinal results, females showed lower amyloid burden and slower accumulation with higher SCFA levels, while the converse was true for males. Our study supports the hypothesis that factors including the gut microbiome could differentially impact AD risk at the earliest disease stages. In contrast, we did not observe that sex modified the effect of SCFA levels on PACC3 trajectories, suggesting that SCFAs may contribute less to sex differences in cognitive decline that previous studies have observed[23]. Together, our cross-sectional and longitudinal results converge on a hypothesis that gut microbiome alterations in midlife and later could reduce gut levels of SCFAs, which may exacerbate amyloid deposition in women and elevate their risk for AD over men.

### 4.4. Potential therapeutic implications

These correlative observations suggest that microbiome-modifying interventions to promote SCFA levels could reduce AD risk. Previous studies have explored this through SCFA supplementation in mice. One APP/PS1 mouse study found that sodium acetate, propionate, and butyrate actually increased amyloid plaque burden and microglial activation[67], possibly because ingested SCFA salts do not reach the large intestine, where microbiota-produced SCFAs may be most protective. However, another study suggests butyrate supplementation could protect against AD, as administering tributyrin, a prodrug of butyrate, prevented hyper-phosphorylation of tau and protected against memory deficits in 3xTg AD mice[19]. Clinical investigations have found tributyrin supplementation to be safe[68], supporting its potential for future trials in AD populations. Propionate supplementation could also be beneficial, as a recent mouse study found that administering sodium propionate to APPPS1-21 mice reduced reactive astrocytosis and amyloid plaques[69]. These studies and our study in humans underscore the need for further investigation to understand the utility of SCFA supplementation in AD.

### 4.5. Strengths and limitations

This study represents the first human study to jointly examine the gut microbiome, SCFAs, and biomarker outcomes in the context of preclinical AD. Strengths include inclusion of cognitively unimpaired A+ individuals, deep characterization of participants’ biomarker, cognitive, and gut microbiome profiles, and longitudinal follow-up on outcome measures. This study also has limitations, including a predominantly White and highly educated cohort from a single study site, potentially limiting generalizability to more diverse populations. Second, there were fewer participants with MCI or dementia due to AD (n=38), and fewer MCI/AD who had CSF biomarker data available (n=8). Lastly, the observational study design yields correlative results, and direction of the effects may be difficult to discern. We cannot rule out the possibility that early brain disease may impact gut microbiome composition or abundance of SCFAs.

In conclusion, these findings support the possibility that SCFAs and their bacterial producers contribute to protection against AD and cognitive decline in humans. This work supports the potential utility of gut-targeted interventions such as SCFA supplementation, probiotics promoting production, or dietary modifications to support SCFA-producing microbes in slowing AD progression.

## Abbreviations

BSS: Bristol Stool Score;
MAGs: metagenome-assembled genomes;
SCFAs: short-chain fatty acids

## Acknowledgements

We thank the WRAP and Wisconsin ADRC participants, as well as the staff at the University of Wisconsin ADRC, the Wisconsin Alzheimer’s Institute, and the Waisman Center for their assistance in study organization, participant recruitment, and data collection. We also thank the University of Wisconsin Biotechnology Center DNA Sequencing Facility for providing sequencing and support services, and the University of Wisconsin Center for High Throughput Computing (CHTC) in the Department of Computer Sciences for providing computational resources, support and assistance. We thank the Landick lab, in particular Bailey Marshall, for use of the Shimadzu GC system and for troubleshooting assistance. We thank Katie Zarbock-Hereford and Matthew F. Warren for training on fecal sample preparation and GC system operation.

The NeuroToolKit is a panel of exploratory prototype assays designed to robustly evaluate biomarkers associated with key pathologic events characteristic of AD and other neurological disorders, used for research purposes only and not approved for clinical use (Roche Diagnostics International Ltd, Rotkreuz, Switzerland). COBAS and ELECSYS are trademarks of Roche. Elecsys β-Amyloid (1–42) CSF, Elecsys Phospho-Tau (181P) CSF and Elecsys Total-Tau CSF assays are approved for clinical use.

## Sources of Funding

NIH R01 AG027161 (the Wisconsin Registry for Alzheimer’s Prevention)

NIH P30 AG062715 (the Wisconsin Alzheimer’s Disease Research Center)

This material is based upon work supported by the National Science Foundation Graduate Research Fellowship Program under Grant No. 2137424. Any opinions, findings, and conclusions or recommendations expressed in this material are those of the author(s) and do not necessarily reflect the views of the National Science Foundation.

Support was also provided by the Graduate School and the Office of the Vice Chancellor for Research at the University of Wisconsin-Madison with funding from the Wisconsin Alumni Research Foundation.

Wisconsin Partnership Program grant (BBB, FER). National Institute on Aging Grants R01AG070973 (BBB, FER, TKU), R01AG083883 (TKU, BBB, FER), R21AG089348 (FER, BBB, TKU). KB is supported by the Swedish Research Council (#2017-00915 and #2022-00732), the Swedish Alzheimer’s Foundation (#AF-930351, #AF-939721, #AF-968270, and #AF-994551), Hjärnfonden, Sweden (#ALZ2022-0006, #FO2024-0048-TK-130 and #FO2024-0048-HK-24), the Swedish state under the agreement between the Swedish government and the County Councils, the ALF-agreement (#ALFGBG-965240 and #ALFGBG-1006418), the European Union Joint Program for Neurodegenerative Disorders (JPND2019-466-236), the Alzheimer’s Association 2021 Zenith Award (ZEN-21-848495), the Alzheimer’s Association 2022-2025 Grant (SG-23-1038904 QC), La Fondation Recherche Alzheimer (FRA), Paris, France, the Kirsten and Freddy Johansen Foundation, Copenhagen, Denmark, Familjen Rönströms Stiftelse, Stockholm, Sweden, and an anonymous philanthropist and donor.

HZ is a Wallenberg Scholar and a Distinguished Professor at the Swedish Research Council supported by grants from the Swedish Research Council (#2023-00356, #2022-01018 and #2019-02397), the European Union’s Horizon Europe research and innovation programme under grant agreement No 101053962, and Swedish State Support for Clinical Research (#ALFGBG-71320).

## Consent Statement

Prior to admission to the study, all participants provided informed consent to be involved in the study.

## Data availability

Shotgun metagenomic data are available from the NCBI Sequence Read Archive (SRA) under accession number PRJNA1271016.

## Author Contributions

Conceptualization: JFK, QZ, MBH, TKU, FER, BBB

Methodology: JFK, QZ, MBH, GK, CQR, BCC, FER, TKU, BBB

Validation: JFK, QZ, MBH

Formal analysis: QZ, MBH

Investigation: JFK, SJH, RLK, VV, ECS, MT, JH, AH, JLW, SS, AM, JZ, HC, EZ, EC, HN, AF, GE, GK, CQR, BCC, FER, TKU, BBB

Resources: CMC, SCJ, SA, HZ, KB, TKU, GK, CQR, BCC, FER, BBB

Data curation: QZ, MBH, JWK, DCP

Writing - Original Draft: JFK, QZ, MBH

Writing - Review & Editing: JFK, QZ, MBH, JWK, SJH, NJDS, DCP, RLK, VV, ECS, MT, JH, AH, JLW, SS, AM, JZ, HC, EZ, EC, HN, AF, GE, GK, CQR, BCC, CMC, SCJ, SA, HZ, KB, TKU, FER, BBB

Visualization: JFK, QZ, MBH

Supervision: SJH, NJDS, FER, BBB

Project administration: JFK, SJH, NJDS, JLW, EC, FER, BBB

Funding acquisition: CMC, SCJ, SA, HZ, KB, BCC, TKU, FER, BBB

