## Supplementary Materials for "Fecal Short-Chain Fatty Acids Vary by Sex and Amyloid Status"

**Methods**

1.1.1. **DNA Extraction**

Fecal DNA was extracted from fecal samples collected from 287 participants using a previously described phenol:chloroform bead-beating protocol [[1]](https://www.zotero.org/google-docs/?21u5Yu). Briefly, feces were subjected to bead-beating twice for three minutes in a mixture containing phenol:chloroform:isoamyl alcohol (UltraPure [25:24:1, v/v], ThermoFisher Scientific). The aqueous phase was collected, and DNA was precipitated by the addition of 3 M sodium acetate and 100% isopropanol. The DNA was then cleaned with the Qiagen/Neucleospin PCR cleanup kit (Macherey-Nagel, Düren, Nordrhein-Westfalen, Germany). DNA concentration was verified using the Qubit dsDNA BR Assay Kit (Life Technologies, Grand Island, NY).

1.1.2. **Metagenomic Sequencing and Processing**

Relative abundances of gut microbial taxa and functional pathways have been measured in fecal samples from 315 MARS participants via shotgun metagenomics with Illumina NovaSeq sequencing. Reads were mapped to taxa and pathways using the metaphlan4 database [[2]](https://www.zotero.org/google-docs/?E9IKmD) and the humann3 database [[3]](https://www.zotero.org/google-docs/?yE3jng).

In 2023-05 batch, 56 samples were sequenced with an average sequencing depth of 55 million pair-end 150 bp reads per sample. In 2024-04 batch, 280 samples were sequenced with an average sequencing depth of 32 million pair-end 150 bp reads per sample. Between 2023-05 batch and 2024-04 batch, there were 21 samples sequenced in both batches. Therefore, there were 315 unique samples sequenced totally, from 294 unique participants.

With the samples analyzed in both batches, we assessed whether species-level abundance of metagenomic features was significantly different across sequencing batches. We used the Bray-Curtis dissimilarity statistic and species present in at least 5% of samples, and determined via visual examination that samples clustered tightly across batch (i.e., that abundances in a given sample were highly similar between batch 1 and 2). Thus, the sequencing reads of these 21 samples were concatenated from 2023 and 2024 batches for downstream analyses.

The microbiome composition phenotypes were generated using MetaPhlAn, on average 67% reads assigned to taxonomy classification. The microbiome pathway phenotypes were generated using HUMAnN. On average, 71% reads mapped to UniRef90, 4.5% reads mapped to Metacyc pathway.

**Shotgun sequencing**

DNA libraries were prepared using Illumina NexteraXT library preparation kit. Quality and quantity of the finished libraries were assessed using an Agilent bioanalyzer and Qubit® dsDNA HS Assay Kit, respectively. Libraries were standardized to 2nM. Paired end, 150 bp sequencing was performed using the Illumina NovaSeq6000, producing 2x150bp paired-end reads. Images were analyzed using the standard Illumina Pipeline (v1.8.2).

**Reads quality control**

Low quality reads were filtered out from raw DNA reads using Trimmomatic (v0.39) with parameters (SLIDINGWINDOW:4:20 MINLEN:50). To identify and eliminate host sequences, reads were aligned against the Homo sapiens genome (GRCh38, Rel109) using bowtie2 (v2.3.4) with default settings, and microbial DNA reads that did not align with the human genome were identified using samtools (v1.3) (samtools view -b -f 4 -f 8).

**Profiling microbiome composition**

Gut microbial taxa were profiled by MetaPhlAn4 pipeline (v4.0.6) using the MetaPhlAn database (mpa_vOct22) and the ChocoPhlAn pan-genome database (mpa_vOct22_CHOCOPhlAnSGB_202212) that contains 5.1M unique clade-specific marker genes identified from ~1M microbial genomes (~236,600 references and 771,500 metagenomic assembled genomes) spanning 26,970 species-level genome bins (SGBs). The unclassified SBG taxa were further annotated to Genome Taxonomy Database (GTDB) using mpa_vOct22_CHOCOPhlAnSGB_202212_SGB2GTDB.tsv data from MetaPhlan4 pipeline. The relative abundances of GTDB taxa were used as microbial composition phenotypes. The GTDB taxa with average relative abundance > 0.01% and present in at least >5% samples were kept for downstream analyses.

**Profiling microbiome functions and pathways**

Gut microbial functions and pathways were profiled using the HUMAnN3 pipeline (v3.7.0), the MetaPhlAn database (mpa_vOct22_CHOCOPhlAnSGB_202212), the ChocoPhlAn pan-genome database (full_chocophlan.v201901_v31) and the UniRef90 protein database (uniref90_annotated_v201901b_full). Pathway abundances were normalized by copy per million (CPM). Uniref90 gene abundances were normalized by copy per million (CPM), then Uniref90 genes were regrouped into KEGG orthology (KO), enzyme commission (EC), gene ontology (GO) and protein family (pfam).

**Metagenome-Assembled Genomes (MAGs)**. Shotgun sequencing reads were assembled for all samples using SPAdes (v.3.15.5; metaspades.py -k 21,33,55,77), the assembled contigs were quantified by mapping shotgun reads to contigs using Bowtie2 (v.2.3.4). Contigs that were less than 500 bp were excluded for downstream analyses. Contigs were then binned into MAGs using MetaBAT2 (v.2.17). The quality of all MAGs was assessed by genome completeness and contamination using CheckM (v.1.2.2) and high-quality MAGs were kept (completeness > 90% and contamination < 5%). The high-quality MAGs were dereplicated using dRep (v.3.5.0; -pa 0.9 -sa 0.99). The final dataset contains 1464 MAGs. To quantify the abundance of each MAG in each sample, shotgun reads from each fecal sample were mapped to MAGs using Bowtie2 (v.2.3.4) and RSEM (v.1.3.1). Taxonomic assignments of these 1464 MAGs were using the Genome Taxonomy Database Toolkit (GTDB-Tk; v.2.3.2) and the GTDB database (ver. R214). Genes from MAGs were predicted using Prodigal (v.2.6.3) and annotated to KEGG Orthology database using kofamscan (v.1.3.0) and annotated to Carbohydrate-Active Enzymes (CAZymes) database using dbcan3 (v.4.1.4) and database “dbCAN3_db_v12_20240415”. MAGs were processed to create a single kallisto quantification index and reads from each faecal DNA sample were mapped to this index to quantify the abundance of each MAG in each sample [[4]](https://www.zotero.org/google-docs/?MDK8W1).

**SCFA gene annotation**. Hidden Markov models (HMM) were built using reference genes from a collection of known propionate-producing bacteria and butyrate-producing bacteria [[5]](https://www.zotero.org/google-docs/?i3fXC0). Reference gene sequences were aligned using MUSCLE (v.3.8.31) [[6]](https://www.zotero.org/google-docs/?oKmXr5), then the HMM profile was constructed using “hmmbuild” from HMMER (v.3.2.1). Protein coding genes (CDS) from MAGs were searched using HMM search (v.3.2.1). All genes involved exhibiting 60% score of the lowest scoring model sequence were included in subsequent analysis. MAGs that have annotated genes methylmalonyl-CoA mutase (mutB), methylmalonyl-CoA epimerase (epi), and methylmalonyl-CoA decarboxylase (mmdA) were identified as propionate producers via the succinate (Suc) pathway; MAGs that have annotated genes propanediol dehydratase (pduCDE) and propionaldehyde dehydrogenase (pduP) were identified as propionate producers via the propanediol (Pdiol) pathway; MAGs that have annotated genes acetyl-CoA acetyltransferase (thl), β-hydroxybutyryl-CoA dehydrogenase (bhbd), crotonase (cro), and butyryl-CoA:acetate CoA transferase (but) were identified as butyrate producers via the Acetyl-CoA pathway.

**Figures**

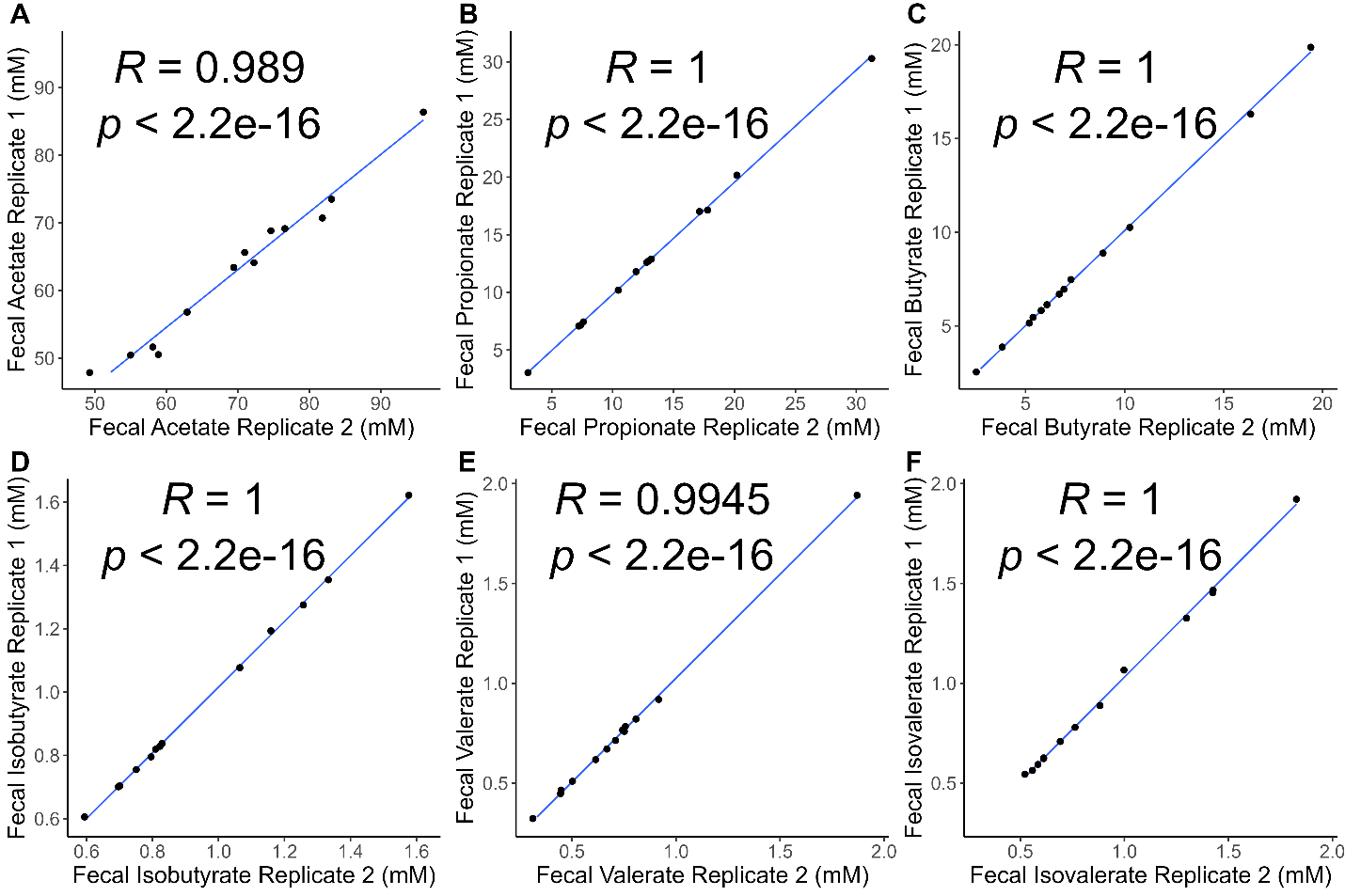

**Supplementary Figure 1. Technical replicates demonstrate high reproducibility of SCFA measurements for fecal A) acetate, B) propionate, C) butyrate, D) isobutyrate, E) valerate, and F) isovalerate.** A subset of 13 samples was randomly selected for repeated measurement within the same analytical run. Spearman’s correlation coefficients (R) were calculated for each SCFA, indicating high reproducibility between technical replicates (R≥0.989, p<2.2e-16).

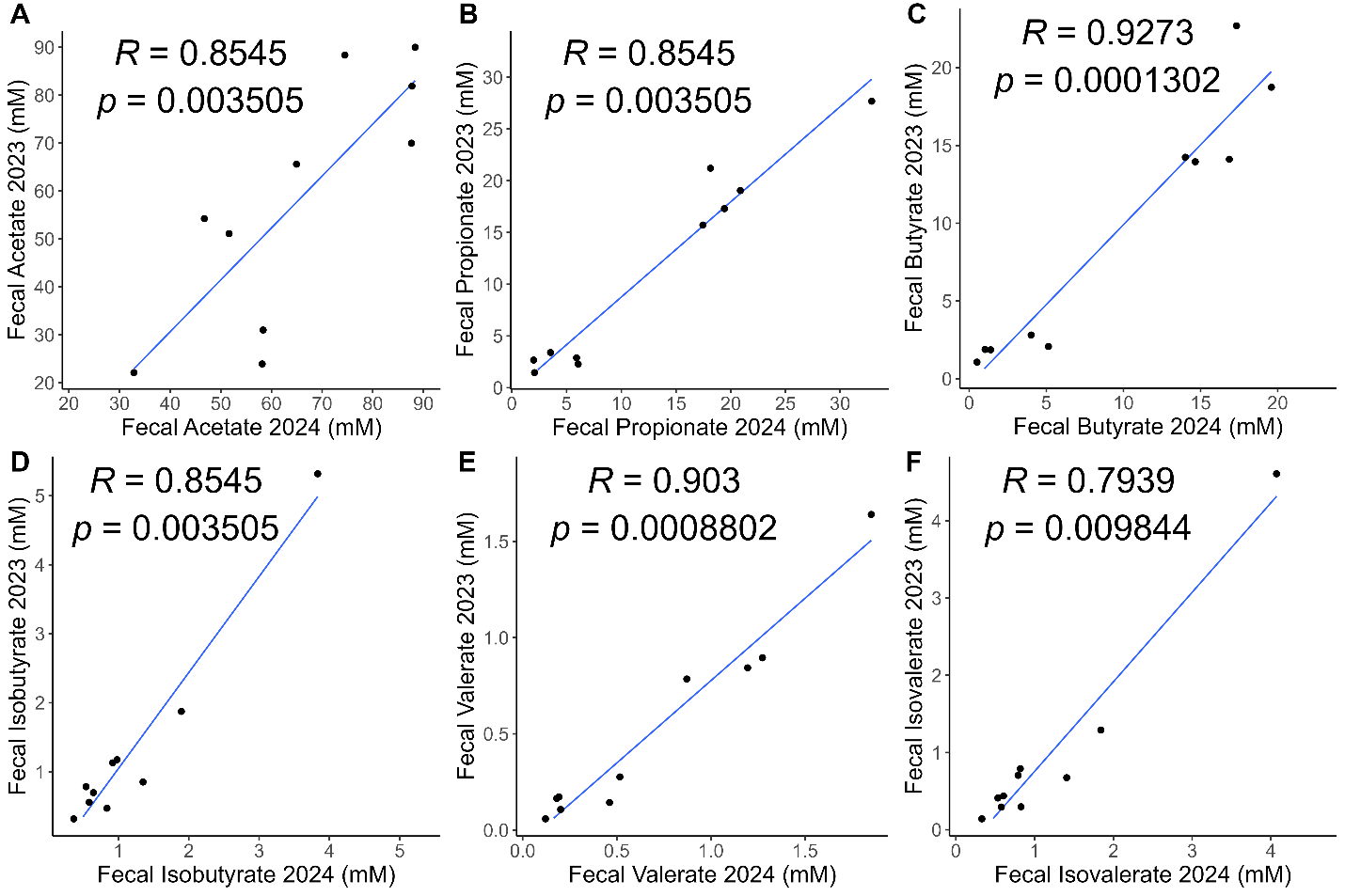

**Supplementary Figure 2. Replicates measured across analytical batches demonstrate high reproducibility of SCFA measurements for fecal A) acetate, B) propionate, C) butyrate, D) isobutyrate, E) valerate, and F) isovalerate.** A subset of 10 samples was randomly selected; for each, two technical replicates were prepared from distinct portions of the same fecal sample and measured in separate batches conducted in different years (2023 and 2024). Spearman’s correlation coefficients (R) were calculated for each SCFA, indicating high reproducibility between technical replicates across batches (R≥0.79, p≤0.003505).

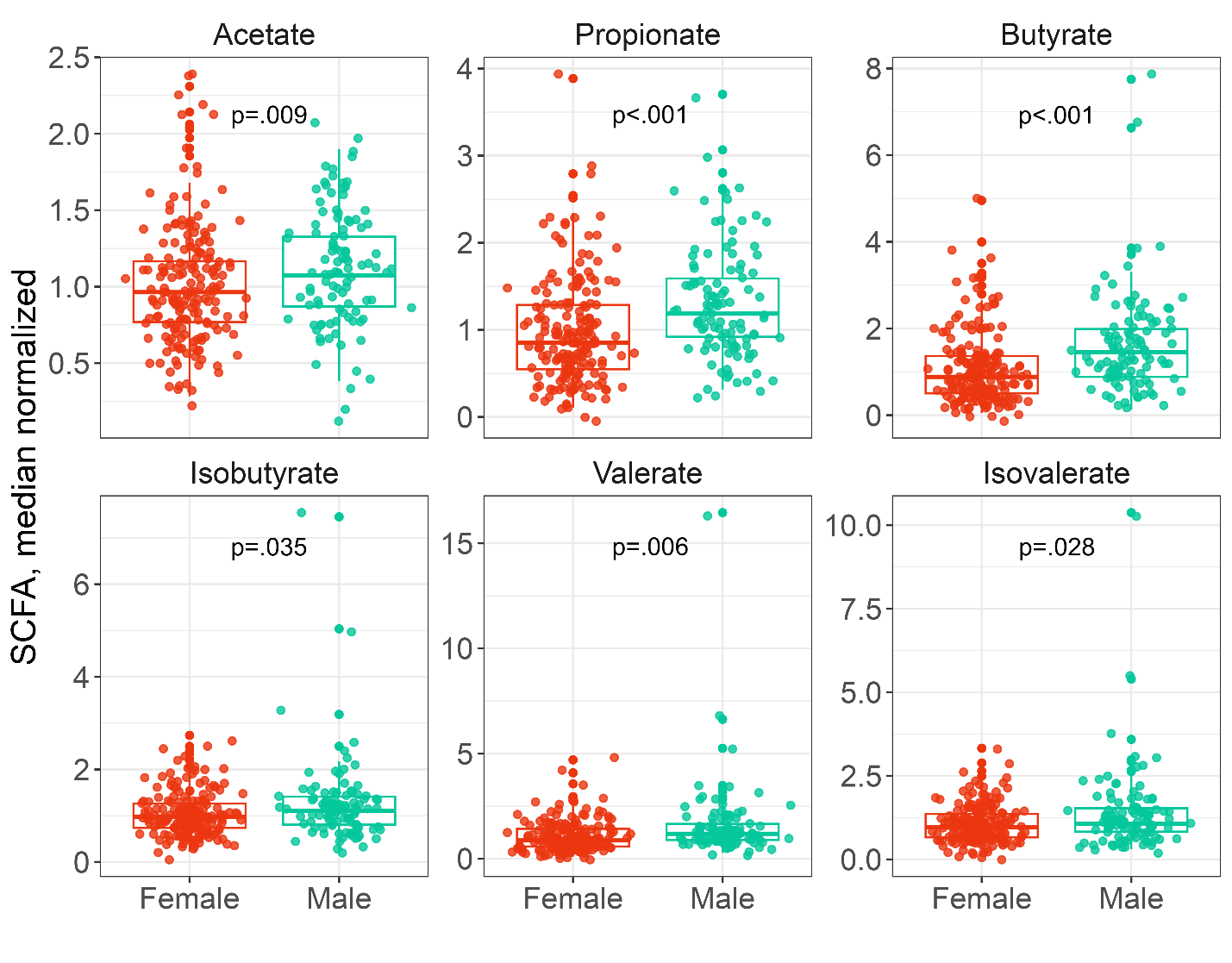

**Supplementary Figure 3. Sex relationships with SCFA abundance.** SCFAs acetate, propionate, butyrate, isobutyrate, valerate, and isovalerate in fecal samples compared across male and female participants using a one-way analysis of means (not assuming equal variances). For each SCFA, abundances are normalized on the median abundance represented in the sample. Abbreviations: SCFA, short-chain fatty acids.

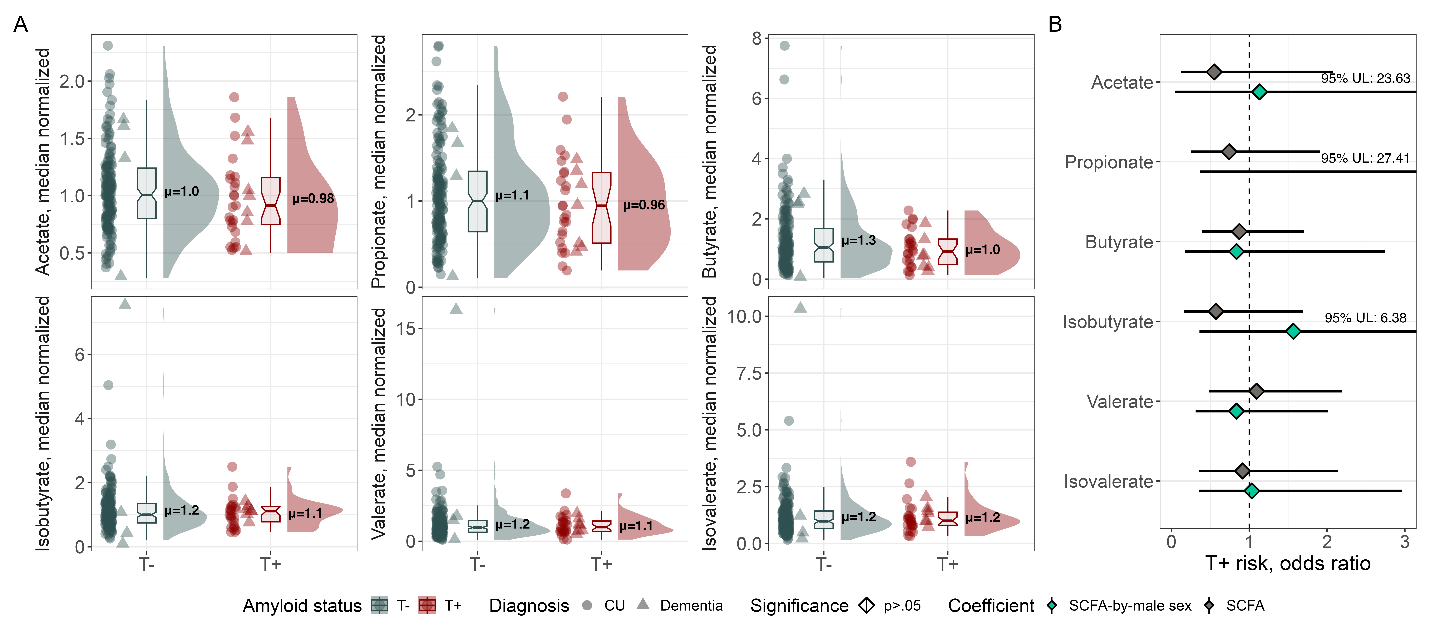

**Supplementary Figure 4. Tau relationships with SCFA abundance.** (**A**) Fecal SCFA levels compared across T+ and T- participants and (**B**) forest plot with odds ratios depicting T+ risk as a function of SCFA level and modified by male sex. From left to right per panel, a scatter plot depicts individual participant values, a box plot shows median SCFA level per tau status, and a density plot shows the data distribution and highlights the mean SCFA level per tau status. SCFA levels in CU participants are depicted with circles, and in MCI or dementia participants with triangles. For each SCFA, abundances are normalized on the median abundance represented in the sample. Odds ratios for main SCFA effects and SCFA-by-sex interactions were derived from the logistic regression model *Tau status ~ SCFA + sex + SCFA-by-sex + age + APOE ε4 carrier status + diagnosis + time between biomarker measurement and fecal collection*. To allow for visual comparison of mean effects, the 95% confidence interval upper limits were annotated for coefficients with wide confidence intervals. Abbreviations: *APOE*, apolipoprotein E; CU, cognitively unimpaired; MCI, mild cognitive impairment; SCFA, short-chain fatty acids; T+/-, tau positive or negative status determined with ^18^F-MK6240 PET; UL, upper limit of the 95% confidence interval.

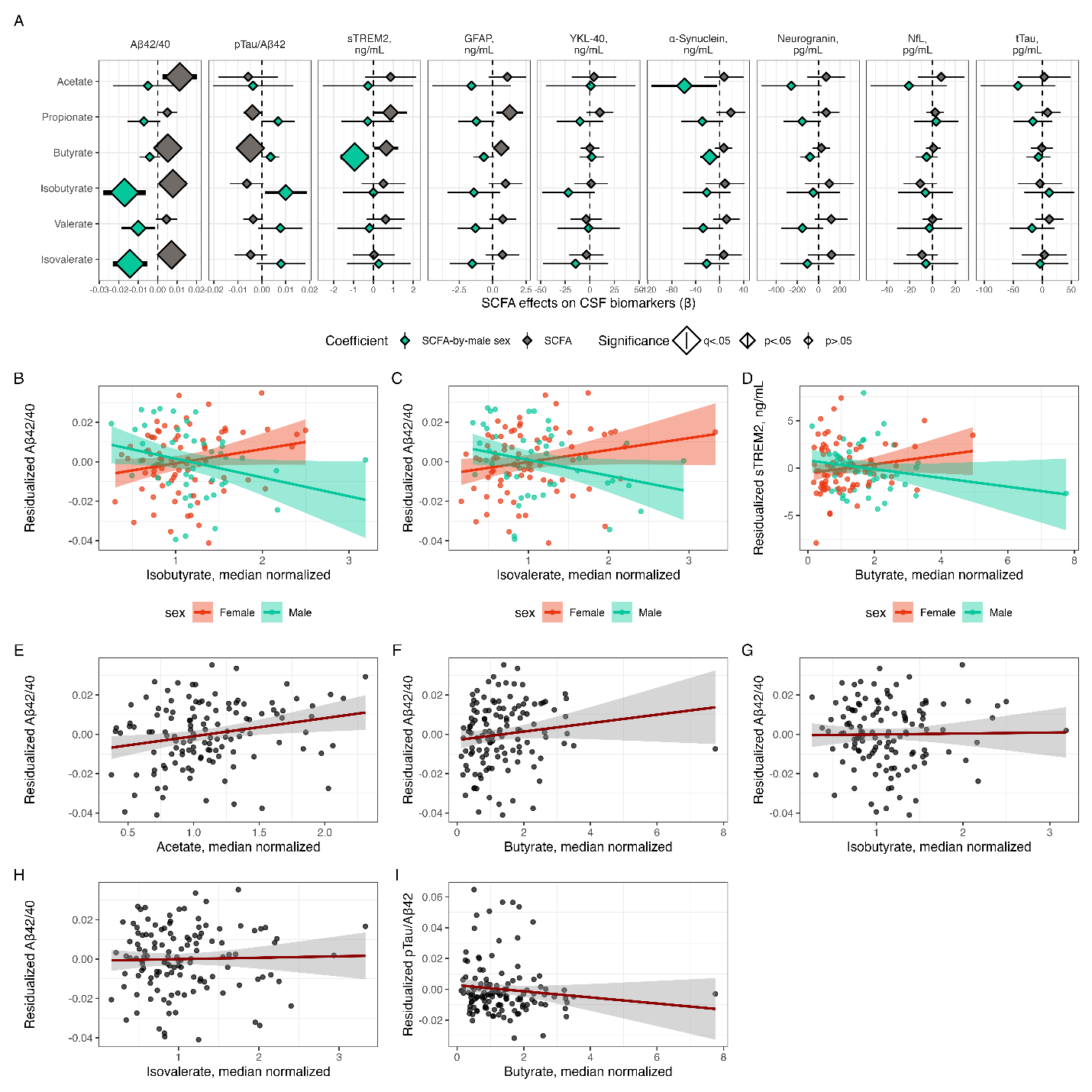

**Supplementary Figure 5. SCFAs significantly associated with CSF biomarkers of amyloid pathology and microglial activation in the full cohort.** (**A**) Forest plot with SCFA and SCFA-by-male sex effect coefficients on CSF biomarker outcomes. Effects were estimated with the regression model *CSF biomarker ~ SCFA + sex + SCFA-by-sex + age + APOE ε4 carrier status + diagnosis + BSS + CSF biomarker batch*. (**B-D**) CSF biomarker levels residualized on model covariates (age, sex, SCFA level, *APOE* ε4 carrier status, diagnosis, BSS, CSF biomarker batch) to demonstrate (**B-C**) Aβ_42_/Aβ_40_ relationships with isobutyrate and isovalerate and (**D**) sTREM2 relationships with butyrate. (**E-I**) Aβ_42_/Aβ_40_ residualized on model covariates (age, sex, *APOE* ε4 carrier status, diagnosis, BSS, CSF biomarker batch) to demonstrate amyloid relationships with (**E**) acetate, (**F**) butyrate, (**G**) isobutyrate, and (**H**) isovalerate main effects. (**I**) pTau_181_/Aβ_42_ residualized on model covariates to demonstrate relationships with butyrate main effects. Abbreviations: Aβ, amyloid-beta; *APOE*, apolipoprotein E; BSS, Bristol Stool Score; GFAP, glial fibrillary acidic protein; NfL, neurofilament light chain protein; pTau_181_, tau phosphorylated at amino acid 181; SCFA, short-chain fatty acids; sTREM2, soluble triggering factor expressed on myeloid cells 2; tTau, total tau; YKL-40, chitinase-3-like protein 1.

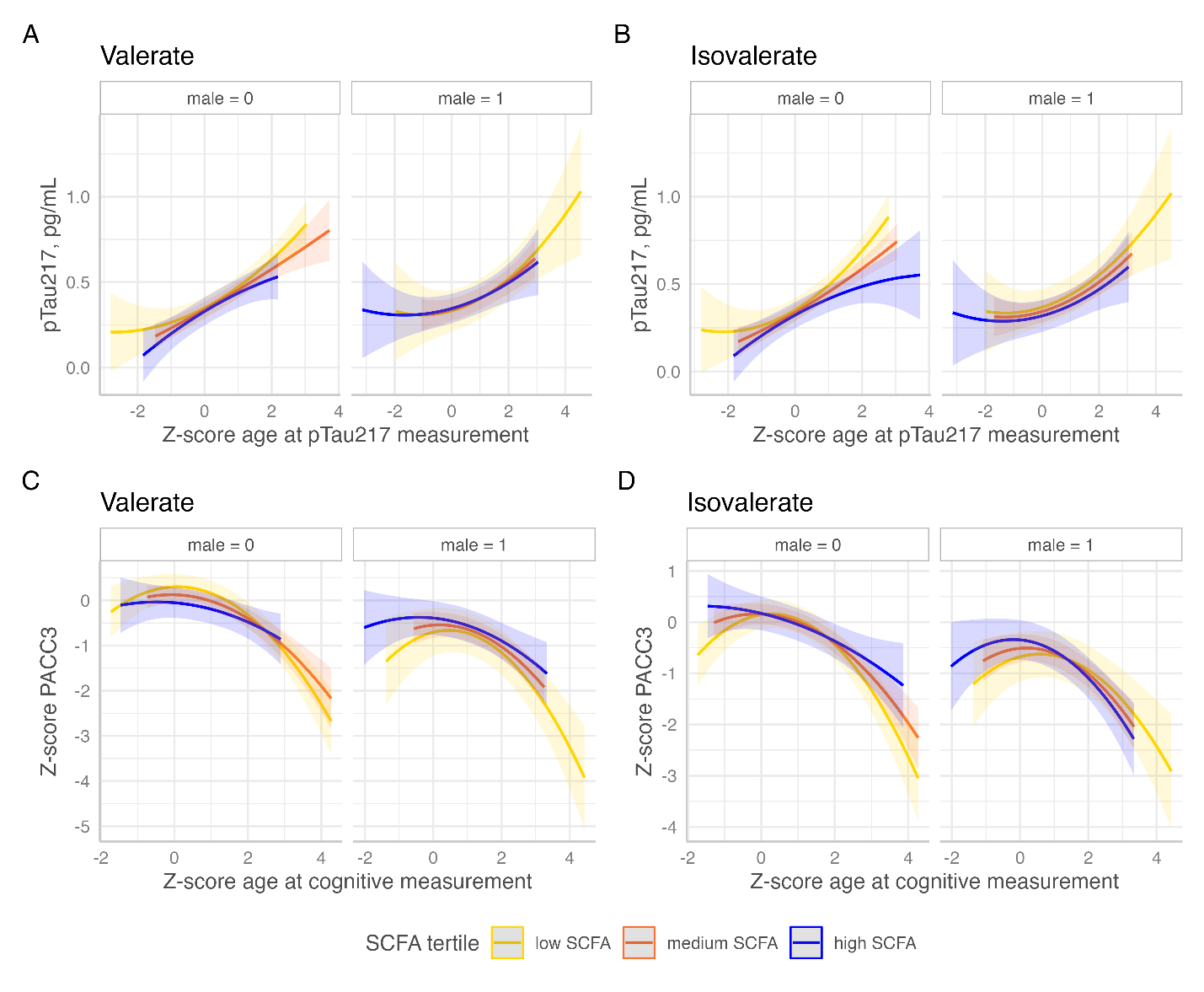

**Supplementary Figure 6. Effects of SCFAs on longitudinal trajectories of pTau_217_ and cognition in the full cohort.** Effects of (**A**,**C**) valerate and (**B**,**D**) isovalerate on longitudinal trajectories of plasma pTau_217_ or cognition (PACC3) by multiple linear regression: *plasma pTau_217_ or cognition ~ SCFA + sex + SCFA-by-male sex + age^2^ + APOE ε4 carrier status + BSS*. For visualization, SCFA levels (normally distributed) are grouped into even tertiles: high, medium, and low. Abbreviations: *APOE*, apolipoprotein E; BSS, Bristol Stool Score; PACC3, Preclinical Alzheimer Cognitive Composite score (three item version); SCFA, short-chain fatty acids.

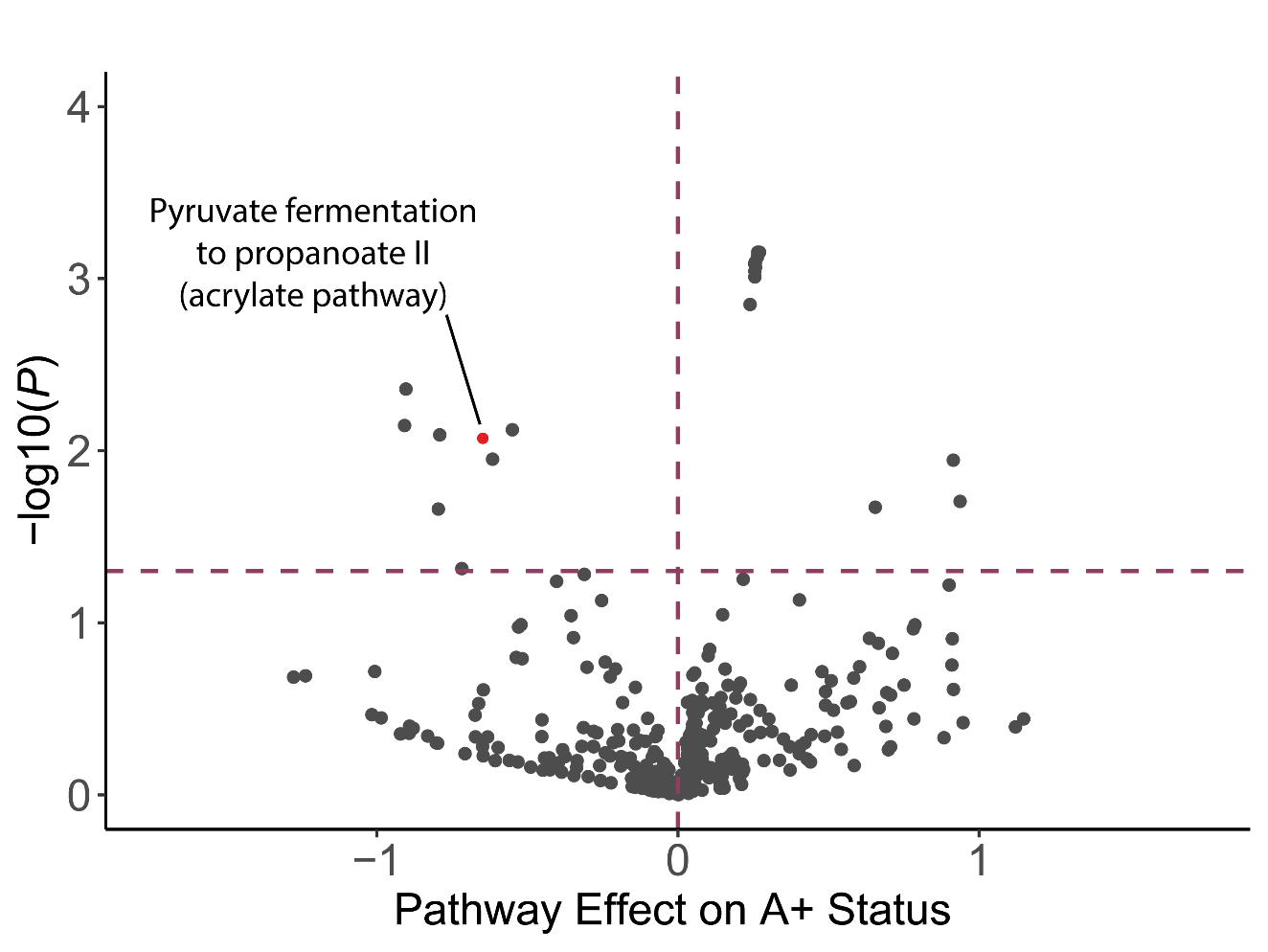

**Supplementary Figure 7. The pyruvate fermentation to propionate MetaCyc pathway was among the most abundant pathways in amyloid-negative participants among cognitively unimpaired participants.** The distribution of log transformed odds ratios for MetaCyc pathways in A+ participants compared to A- (negative values = A-, positive values = A+). Effects of MetaCyc pathways on amyloid status were assessed by logistic regression: *Amyloid status ~ MetaCyc pathway + sex + age + APOE ε4 carrier status + BSS.* These associations did not survive FDR correction (pyruvate fermentation to propionate pathway: p=0.008, q=0.206). Abbreviations: A+/-, amyloid positive or negative status determined with ^11^C-PiB PET, CSF Aβ_42_/Aβ_40_, or plasma pTau_217_; *APOE*, apolipoprotein E; BSS, Bristol Stool Score.

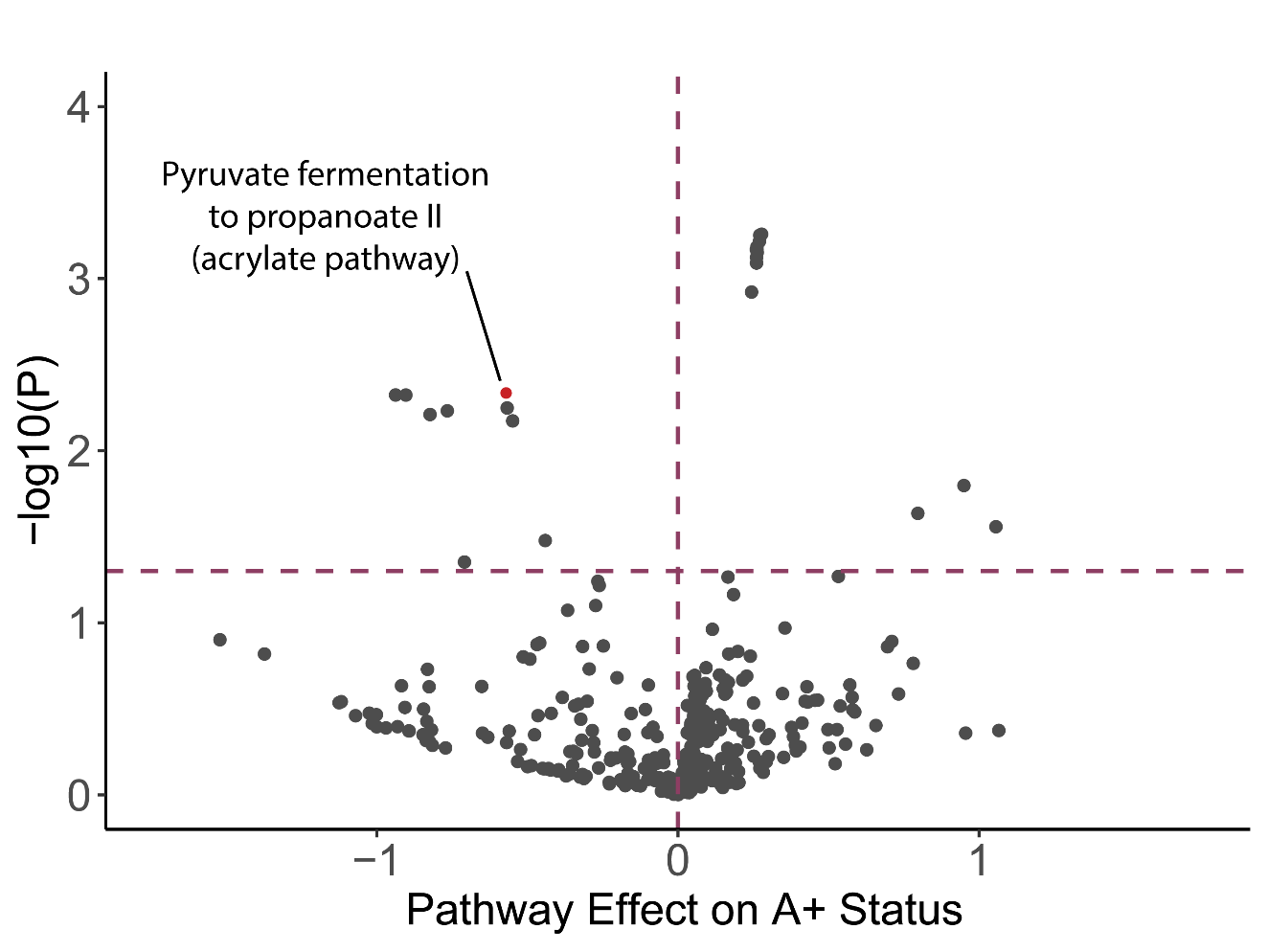

**Supplementary Figure 8. The pyruvate fermentation to propionate MetaCyc pathway was among the most abundant pathways in amyloid-negative participants among the full cohort.** The distribution of log transformed odds ratios for MetaCyc pathways in A+ participants compared to A- (negative values = A-, positive values = A+). Effects of MetaCyc pathways on amyloid status were assessed by logistic regression: *Amyloid status ~ MetaCyc pathway + sex + age + APOE ε4 carrier status + BSS + diagnosis.* These associations did not survive FDR correction (pyruvate fermentation to propionate pathway: p=0.005, q=0.125). Abbreviations: A+/-, amyloid positive or negative status determined with ^11^C-PiB PET, CSF Aβ_42_/Aβ_40_, or plasma pTau_217_; *APOE*, apolipoprotein E; BSS, Bristol Stool Score.

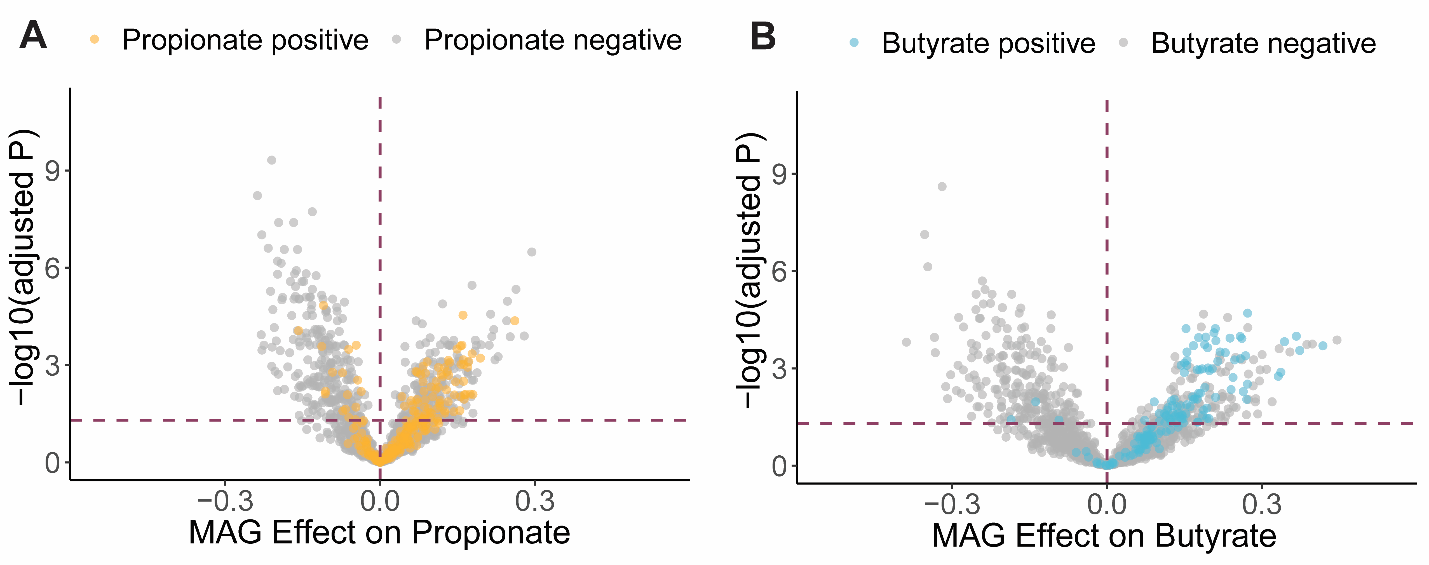

**Supplementary Figure 9.** **MAGs with A) propionate and B) butyrate producing genes have higher relative abundances in samples with high levels of fecal propionate and butyrate, respectively, in the cognitively unimpaired cohort.** Effect coefficients for MAGs with genetic pathways for (**A**) propionate production and (**B**) butyrate production in participants with high levels of fecal propionate or butyrate compared to low levels of fecal propionate or butyrate, respectively, for producer MAGs vs. non-producer MAGs (labeled propionate or butyrate positive or negative by color). Effects of MAGs on SCFA levels (negative values = low SCFA levels, positive values = high SCFA levels) were estimated with the regression model *SCFA ~ MAG + age + sex + BSS*. Propionate positive and Butyrate positive MAGs were classified based on gene content. Abbreviations: BSS, Bristol Stool Score; SCFA, short-chain fatty acid; MAG, metagenome-assembled genome.

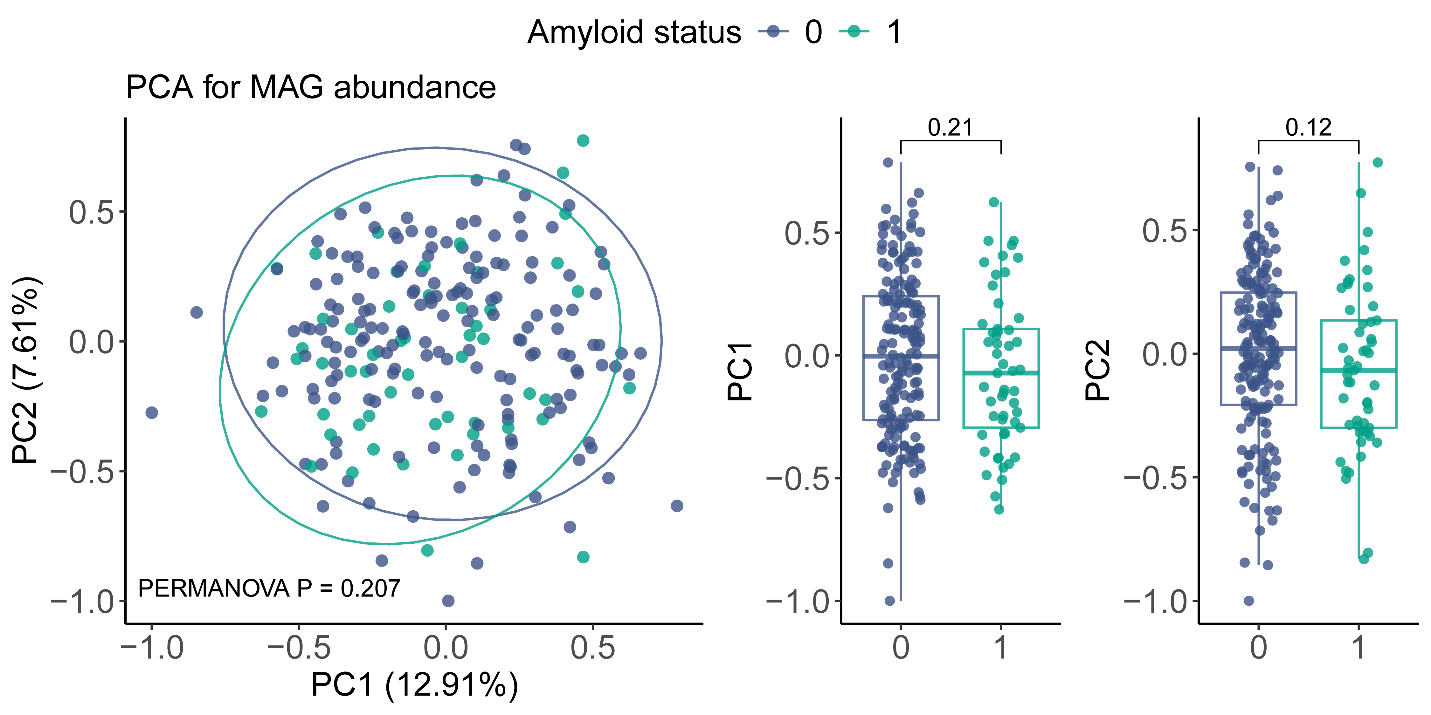

**Supplementary Figure 10. PERMANOVA with Bray-Curtis dissimilarity was used to assess whether gut microbiome features (MAGs) are significantly different between A- and A+ participants in the cognitively unimpaired cohort.** MAG relative abundances were centered log ratio transformed. PC1 and PC2 axes represent the most variance in the data. PERMANOVA was used to determine statistical significance. Abbreviations: PC1, principal component 1; PC2, principal component 2. Abbreviations: MAG, metagenome-assembled genome.

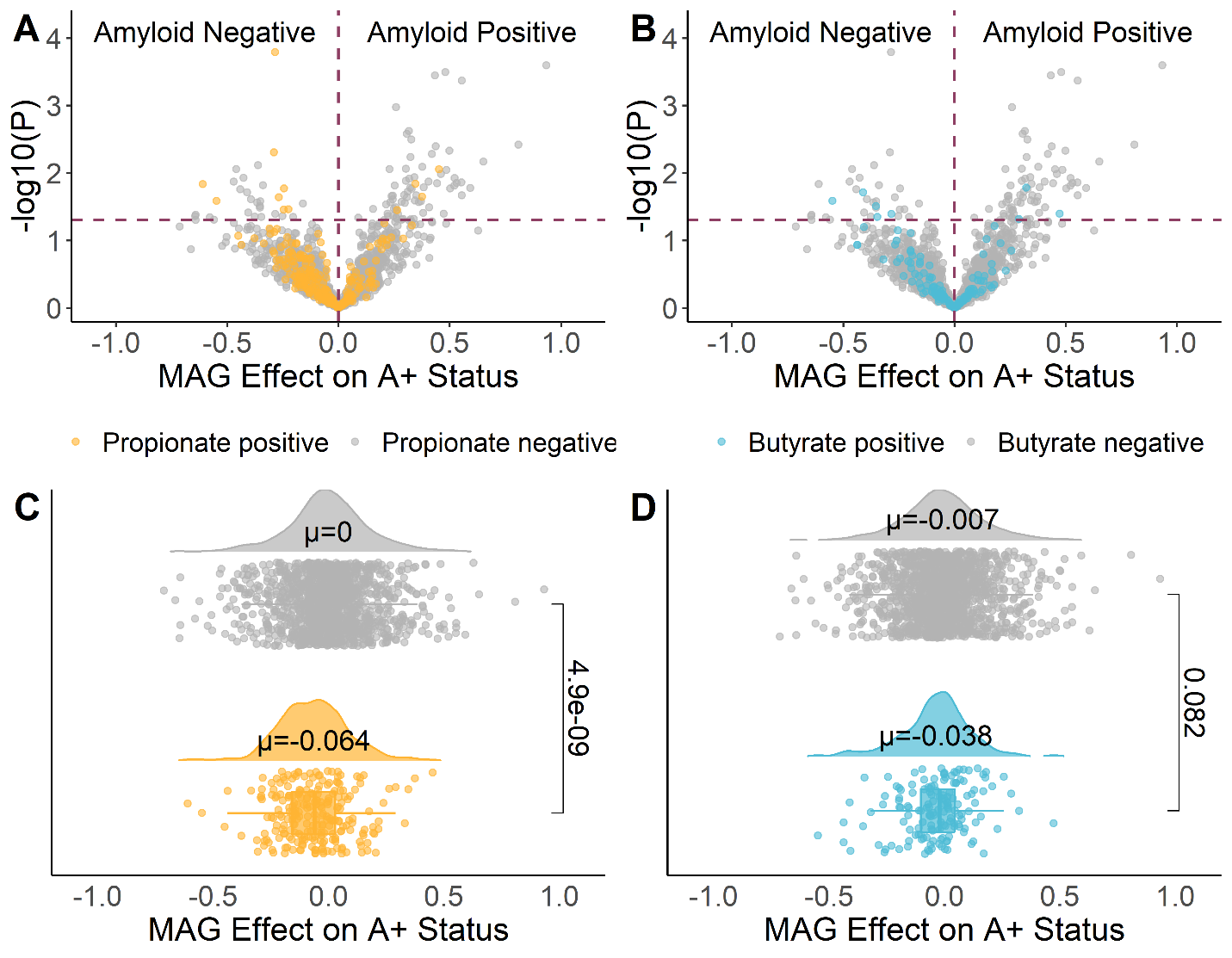

**Supplementary Figure 11. Distributions of propionate- and butyrate-producing bacteria are shifted toward amyloid-negative status in the full cohort.** The distribution of log transformed odds ratios (μ) for MAGs with genetic pathways for (**A**,**C**) propionate production and (**B**,**D**) butyrate production in A+ participants compared to A-, for producer MAGs vs. non-producer MAGs (labeled propionate or butyrate positive or negative by color), as assessed by a Wilcoxon rank-sum test for log transformed odds ratios, indicating the effect of each MAG on amyloid status (negative values = A-, positive values = A+). Effects of MAGs on amyloid status were assessed by logistic regression: *Amyloid status ~ MAG + sex + age + APOE ε4 carrier status + BSS.* (**C**,**D**): From top to bottom per panel, a density plot shows the data distribution, highlighting the mean MAG log transformed odds ratio per group, and a scatter plot depicts the log transformed odds ratio for each MAG. Propionate Positive and Butyrate Positive MAGs were classified based on gene content. Abbreviations: A+/-, amyloid positive or negative status determined with ^11^C-PiB PET, CSF Aβ_42_/Aβ_40_, or plasma pTau_217_; *APOE*, apolipoprotein E; BSS, Bristol Stool Score; MAG, metagenome-assembled genome.

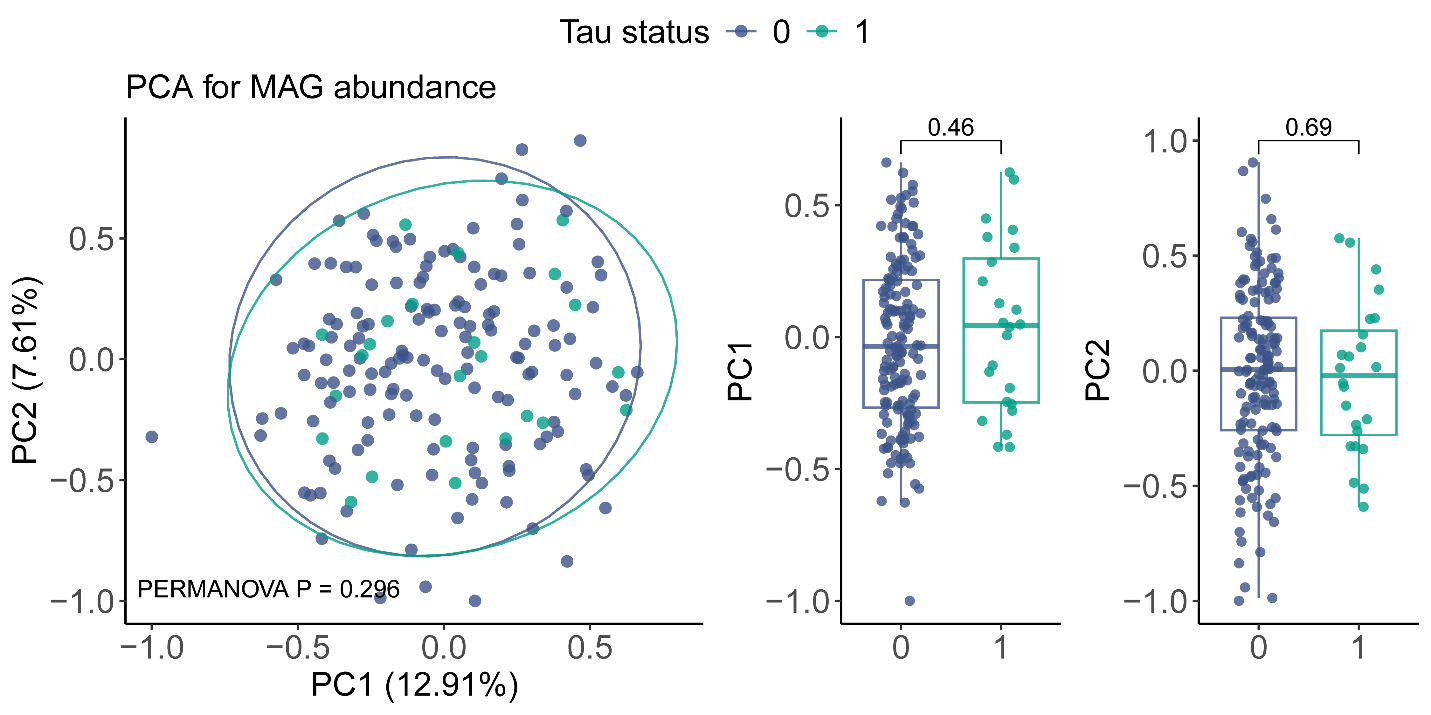

**Supplementary Figure 12. PERMANOVA with bray-curtis dissimilarity was used to assess whether gut microbiome features (MAGs) are significantly different between T- and T+ participants in the cognitively unimpaired cohort.** MAG relative abundances were centered log ratio transformed. PC1 and PC2 axes represent the most variance in the data. PERMANOVA was used to determine statistical significance. Abbreviations: PC1, principal component 1; PC2, principal component 2; T+/-, tau positive or negative status determined with ^18^F-MK6240 PET; MAG, metagenome-assembled genome.

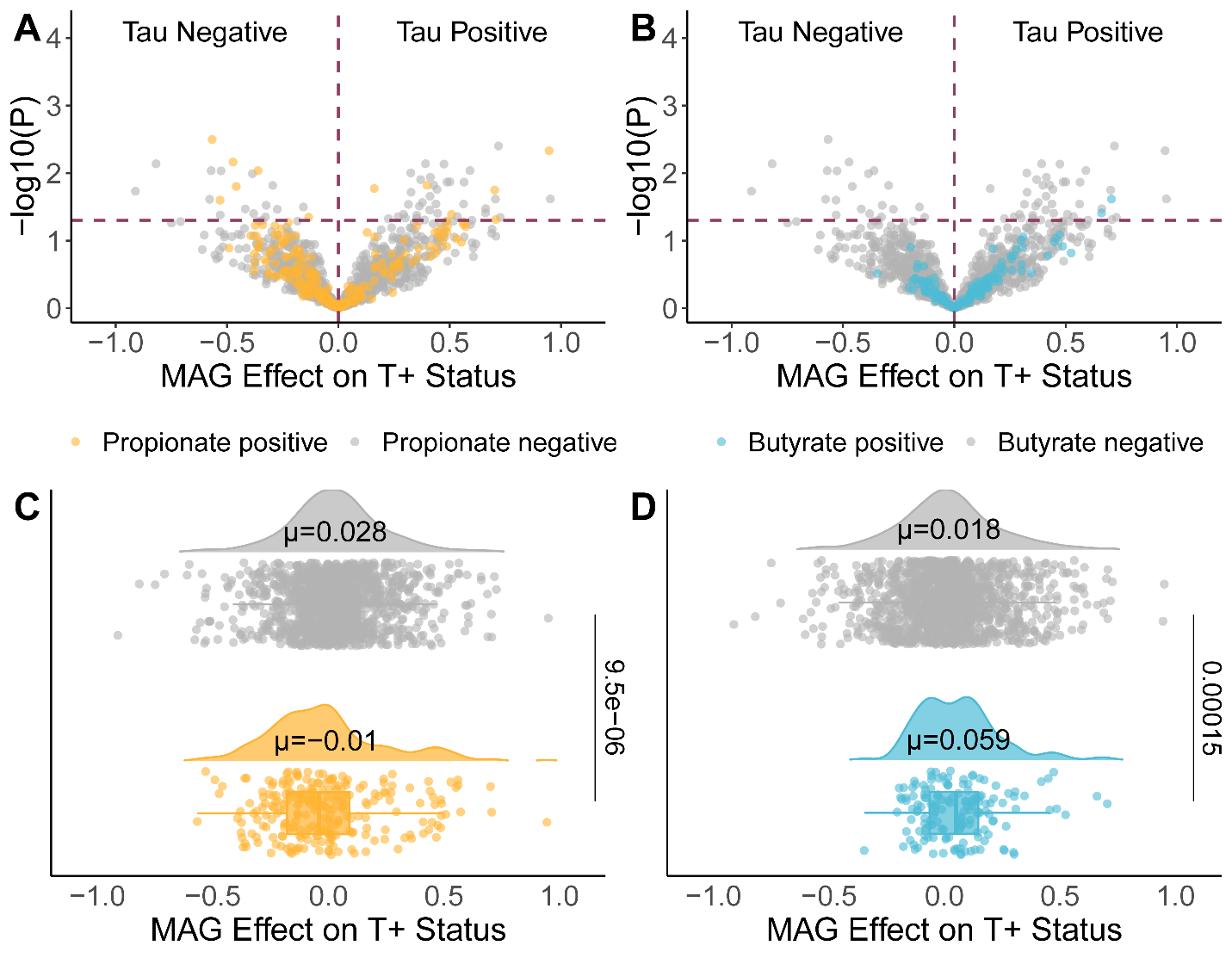

**Supplementary Figure 13. Distributions of propionate- and butyrate-producing bacteria among cognitively unimpaired participants are shifted toward tau-negative status.** The distribution of log transformed odds ratios (μ) for MAGs with genetic pathways for (**A**,**C**) propionate production and (**B**,**D**) butyrate production in T+ participants compared to T-, for producer MAGs vs. non-producer MAGs (labeled propionate or butyrate positive or negative by color), as assessed by a Wilcoxon rank-sum test for ꞵ coefficients, indicating the effect of each MAG on tau status (negative values = T-, positive values = T+). Effects of MAGs on tau status were assessed by logistic regression: *Tau status ~ MAG + sex + age + APOE ε4 carrier status + BSS.* (**C**,**D**): From top to bottom per panel, a density plot shows the data distribution, highlighting the mean MAG log transformed odds ratio per group, and a scatter plot depicts the log transformed odds ratio for each MAG. Propionate Positive and Butyrate Positive MAGs were classified based on gene content. Abbreviations: *APOE*, apolipoprotein E; BSS, Bristol Stool Score; T+/-, tau positive or negative status determined with ^18^F-MK6240 PET; MAG, metagenome-assembled genome.

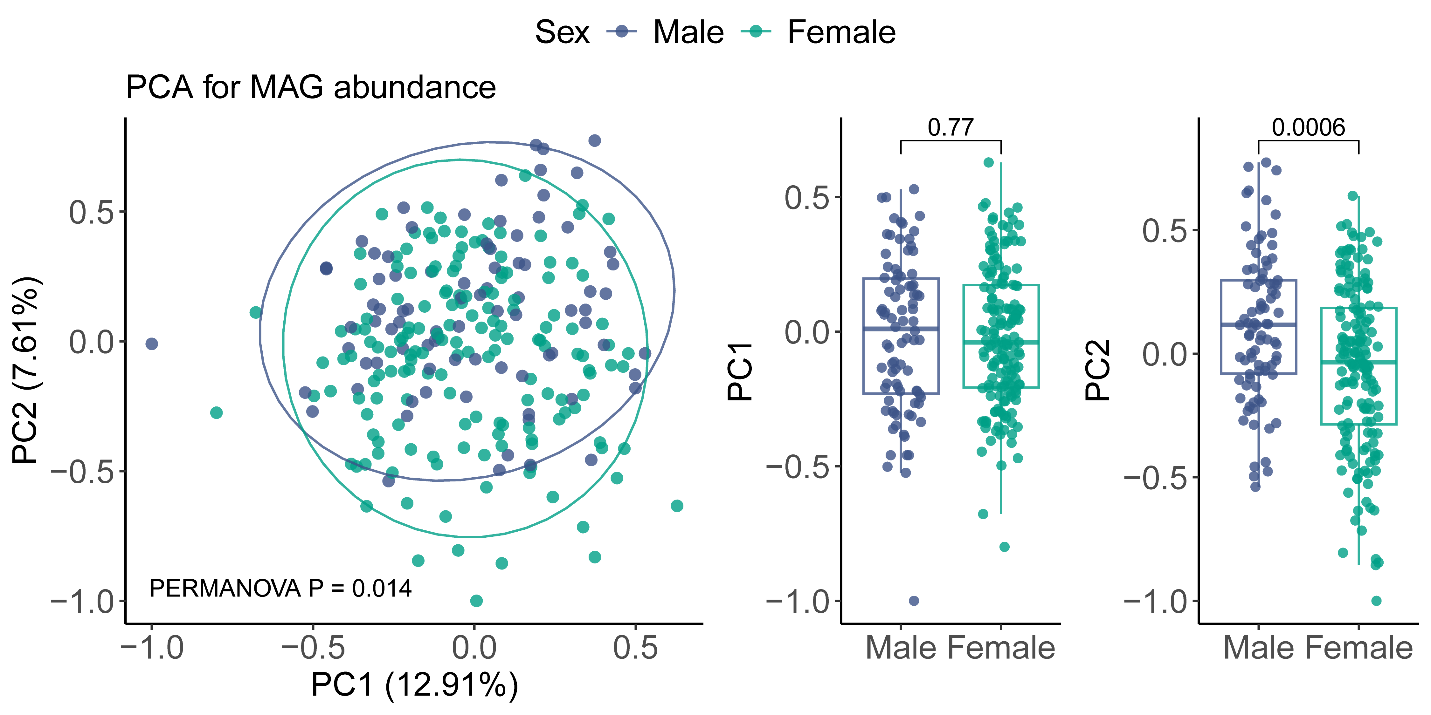

**Supplementary Figure 14. PERMANOVA with bray-curtis dissimilarity was used to assess whether gut microbiome features (MAGs) are significantly different between males and females in the cognitively unimpaired cohort.** MAG relative abundances were centered log ratio transformed. PC1 and PC2 axes represent the most variance in the data. PERMANOVA was used to determine statistical significance. Abbreviations: PC1, principal component 1; PC2, principal component 2; MAG, metagenome-assembled genome.

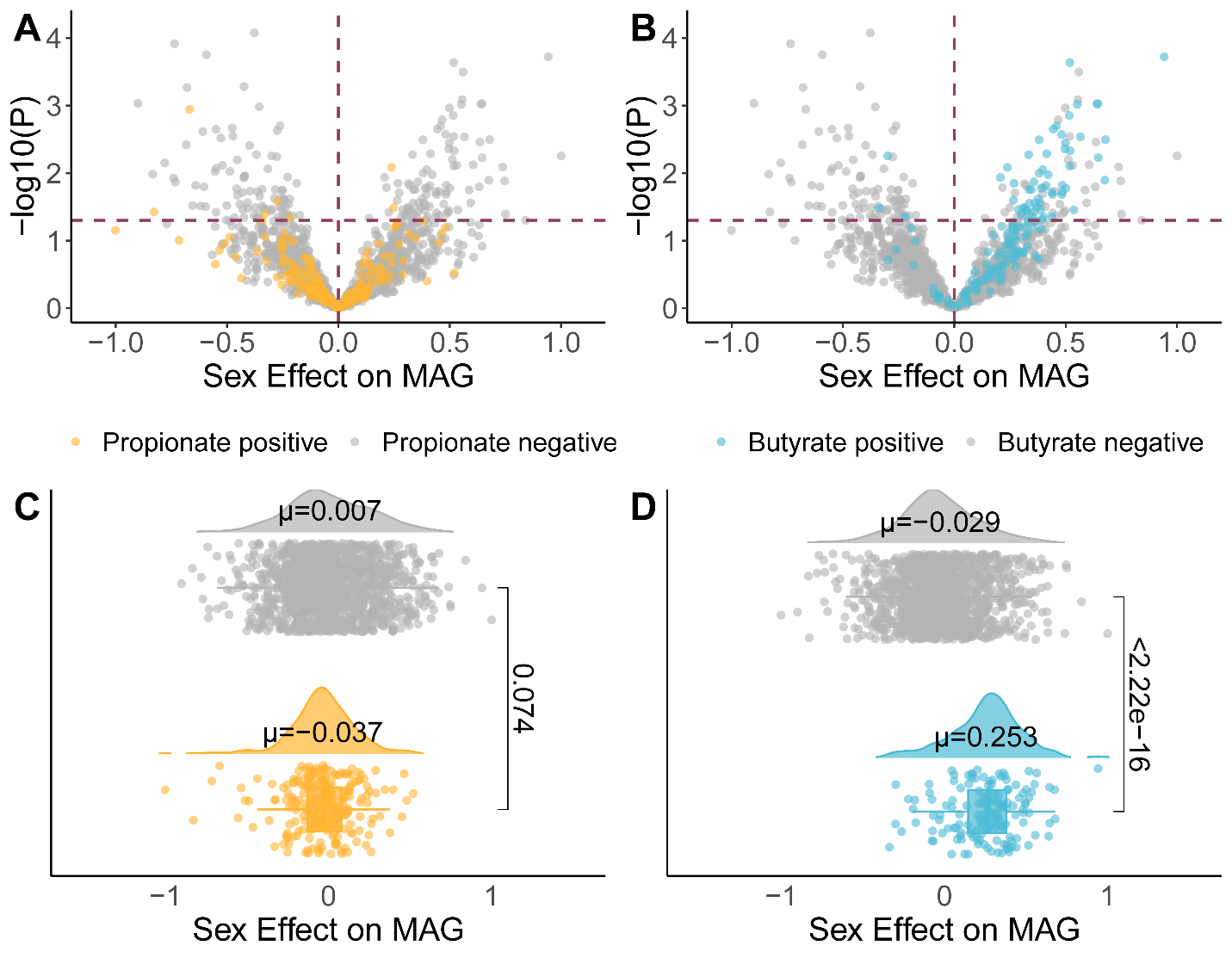

**Supplementary Figure 15. Males have a significantly higher abundance of butyrate-producing MAGs than females.** The distribution of effect coefficients for MAGs with genetic pathways for (**A**,**C**) propionate production and (**B**,**D**) butyrate production in male participants compared to females, for producer MAGs vs. non-producer MAGs (labeled propionate or butyrate positive or negative by color), as assessed by a Wilcoxon rank-sum test for effect coefficients, indicating the effect of each sex on MAGs (negative values = females, positive values = males). Effects of sex on MAGs were estimated with the regression model *MAG ~ Sex + age + BSS.* (**C**,**D**): From top to bottom per panel, a density plot shows the data distribution, highlighting the mean MAG effect coefficient per group, and a scatter plot depicts the effect coefficient for each MAG. Propionate Positive and Butyrate Positive MAGs were classified based on gene content. Abbreviations: BSS, Bristol Stool Score; MAG, metagenome-assembled genome.

**Tables**

**Supplementary Table 1. Participant medication usage.**

| **Characteristic** | **Overall**   N = 286* | **AD**   N = 38* | **CU**   N = 24* | **CU A-**   N = 171* | **CU A+**   N = 53* | ***P* value** |
| --- | --- | --- | --- | --- | --- | --- |
| ACE inhibitors | 49 (17) | 10 (26) | 4 (17) | 28 (16) | 7 (13) | 0.4^✝^ |
| Alpha Blockers | 17 (5.9) | 4 (11) | 2 (8.3) | 8 (4.7) | 3 (5.7) | 0.4^✝^ |
| Analgesics | 28 (9.8) | 5 (13) | 4 (17) | 14 (8.2) | 5 (9.4) | 0.4^✝^ |
| Angiotensin II Receptor Blockers | 23 (8.0) | 5 (13) | 2 (8.3) | 12 (7.0) | 4 (7.5) | 0.6^✝^ |
| Anti Platelets (Aspirin) | 108 (38) | 16 (42) | 11 (46) | 62 (36) | 19 (36) | 0.7^☨^ |
| Antibiotics | 13 (4.5) | 1 (2.6) | 0 (0) | 10 (5.8) | 2 (3.8) | 0.8^✝^ |
| Anticholinesterases | 20 (7.0) | 20 (53) | 0 (0) | 0 (0) | 0 (0) | <0.001^✝^ |
| Anticonvulsants | 15 (5.2) | 5 (13) | 1 (4.2) | 9 (5.3) | 0 (0) | 0.038^✝^ |
| Antidepressants |  |  |  |  |  |  |
| SNRIs | 14 (4.9) | 1 (2.6) | 1 (4.2) | 9 (5.3) | 3 (5.7) | >0.9^✝^ |
| SSRIs | 55 (19) | 16 (42) | 1 (4.2) | 28 (16) | 10 (19) | 0.001^✝^ |
| TCA Antidepressants | 8 (2.8) | 0 (0) | 1 (4.2) | 6 (3.5) | 1 (1.9) | 0.7^✝^ |
| Other Antidepressants | 26 (9.1) | 7 (18) | 0 (0) | 13 (7.6) | 6 (11) | 0.066^✝^ |
| Anti-anxiolytics  Benzodiazepines | 14 (4.9) | 3 (7.9) | 1 (4.2) | 9 (5.3) | 1 (1.9) | 0.6^✝^ |
| Non-benzodiazepines | 7 (2.4) | 0 (0) | 0 (0) | 5 (2.9) | 2 (3.8) | 0.7^✝^ |
| Atypical antipsychotics | 2 (0.7) | 2 (5.3) | 0 (0) | 0 (0) | 0 (0) | 0.024^✝^ |
| Beta Blockers | 37 (13) | 7 (18) | 5 (21) | 19 (11) | 6 (11) | 0.3^✝^ |
| Blood glucose lowering drugs | 26 (9.1) | 4 (11) | 3 (13) | 16 (9.4) | 3 (5.7) | 0.7^✝^ |
| Bronchodilators | 30 (10) | 3 (7.9) | 4 (17) | 19 (11) | 4 (7.5) | 0.6^✝^ |
| Calcium Channel Blockers | 23 (8.0) | 5 (13) | 4 (17) | 11 (6.4) | 3 (5.7) | 0.2^✝^ |
| Cholesterol Lowering (Non-Statin) | 7 (2.4) | 1 (2.6) | 0 (0) | 3 (1.8) | 3 (5.7) | 0.3^✝^ |
| Corticosteroids | 14 (4.9) | 3 (7.9) | 1 (4.2) | 8 (4.7) | 2 (3.8) | 0.8^✝^ |
| Diuretics  Gastrointestinal | 49 (17) | 7 (18) | 4 (17) | 29 (17) | 9 (17) | >0.9^✝^ |
| Antacids | 8 (2.8) | 0 (0) | 0 (0) | 7 (4.1) | 1 (1.9) | 0.7^✝^ |
| H2 Blockers | 9 (3.1) | 2 (5.3) | 1 (4.2) | 4 (2.3) | 2 (3.8) | 0.5^✝^ |
| PPIs | 38 (13) | 6 (16) | 2 (8.3) | 22 (13) | 8 (15) | 0.8^✝^ |
| Hormone Replacements | 25 (8.7) | 0 (0) | 1 (4.2) | 19 (11) | 5 (9.4) | 0.11^✝^ |
| Laxatives | 21 (7.3) | 5 (13) | 1 (4.2) | 13 (7.6) | 2 (3.8) | 0.4^✝^ |
| Memantine | 13 (4.5) | 13 (34) | 0 (0) | 0 (0) | 0 (0) | <0.001^✝^ |
| Muscle Relaxants | 9 (3.1) | 0 (0) | 1 (4.2) | 7 (4.1) | 1 (1.9) | 0.6^✝^ |
| NSAIDs | 62 (22) | 5 (13) | 6 (25) | 40 (23) | 11 (21) | 0.6^☨^ |
| Ophthalmics | 21 (7.3) | 1 (2.6) | 3 (13) | 14 (8.2) | 3 (5.7) | 0.5^✝^ |
| Probiotics | 10 (3.5) | 0 (0) | 0 (0) | 8 (4.7) | 2 (3.8) | 0.6^✝^ |
| Statins | 124 (43) | 26 (68) | 11 (46) | 66 (39) | 21 (40) | 0.009^✝^ |
| Thyroid hormones  Vitamins and Supplements | 65 (23) | 13 (34) | 3 (13) | 39 (23) | 10 (19) | 0.2^☨^ |
| Calcium | 63 (22) | 7 (18) | 4 (17) | 39 (23) | 13 (25) | 0.8^☨^ |
| Melatonin | 16 (5.6) | 2 (5.3) | 1 (4.2) | 12 (7.0) | 1 (1.9) | 0.6^✝^ |
| Multivitamins | 116 (41) | 17 (45) | 12 (50) | 66 (39) | 21 (40) | 0.7^☨^ |
| Omega 3 triglycerides | 63 (22) | 9 (24) | 6 (25) | 42 (25) | 6 (11) | 0.2^☨^ |
| Vitamin B | 40 (14) | 8 (21) | 0 (0) | 24 (14) | 8 (15) | 0.094^✝^ |
| Vitamin C | 26 (9.1) | 2 (5.3) | 0 (0) | 17 (9.9) | 7 (13) | 0.2^✝^ |
| Vitamin D & analogues | 170 (59) | 18 (47) | 11 (46) | 106 (62) | 35 (66) | 0.14^☨^ |
| Vitamin E | 16 (5.6) | 6 (16) | 0 (0) | 7 (4.1) | 3 (5.7) | 0.040^✝^ |

*n (%) ^✝^Fisher's exact test ^☨^Pearson's Chi-squared test. Medications related to the gut microbiome or Alzheimer’s disease, or reported by more than 10 participants. Medication data was collected at the study visit closest to fecal sample collection, and was unavailable for one participant. Blood glucose lowering drugs include biguanides (metformin), insulin, GLP-1 receptor agonists, SGLT2 inhibitors (Jardiance), DPP-4 inhibitors (Tradjenta), and sulfonylureas (glimepiride).

**Supplementary Table 2. Preanalytic protocols for plasma biomarker measurement.**

| **Dates** | **Collection Tube** | **Inversion** | **Centrifugation** | **Storage Tube** | **Storage Temperature** |
| --- | --- | --- | --- | --- | --- |
| 2011-2020 | 9 mL heparin | 8-10 times after blood draw  8-10 times before centrifuging | 15 minutes at 2000 g and 4^°^C | 0.5 mL aliquots  1.5 mL microcentrifuge tubes (2011-2020) or 1.0 mL matrix cryotubes (2015-2019) | -70^°^C  Transitioned to -80^°^C in 2013 |
| 2020-2021 | 6 mL K_2_ EDTA |  | 10 minutes at 2000 g and 4^°^C | 0.5 mL aliquots  1.5 mL microcentrifuge tubes | -80^°^C |
| 2021-2022 |  |  |  | 0.5 mL aliquots  0.5 Sarstedt cryotubes | -80^°^C  Frozen within 1 hour of collection |
| 2022-2023 |  | 8-10 times after blood draw |  |  |  |

Cells that span multiple rows are used to denote procedures that were sustained across time periods (iterations of the preanalytic protocol).

**Supplementary Table 3. Participant demographics compared across analysis subsets.**

| **Characteristic** | **Full cohort**  N = 287 | **Amyloid status**  **Goals 1 and 3**  N = 242 | **Tau status**  **Goal 1**  N = 186 | **CSF biomarkers**  **Goals 1 and 3**  N = 127 | ***P* value** |
| --- | --- | --- | --- | --- | --- |
| Clinical diagnosis, n (%) |  |  |  |  | 0.012* |
| Cognitively unimpaired | 249 (87) | 225 (93) | 175 (94) | 119 (94) |  |
| Dementia | 38 (13) | 17 (7.0) | 11 (5.9) | 8 (6.3) |  |
| A+ at fecal collection, n (%) | 65 (27) | 65 (27) | 53 (29) | 39 (31) | 0.8* |
| Unknown amyloid status, n | 45 | 0 | 3 | 1 |  |
| T+ at fecal collection, n (%) | 31 (17) | 31 (17) | 31 (17) | 15 (17) | >0.9* |
| Unknown tau status, n | 101 | 59 | 0 | 38 |  |
| Age at fecal collection, mean (SD) years | 68 (7.1) | 67 (6.7) | 68 (6.6) | 66 (6.6) | 0.3^☨^ |
| Sex, n (%) |  |  |  |  | 0.8* |
| Female | 184 (64) | 157 (65) | 125 (67) | 78 (61) |  |
| Male | 103 (36) | 85 (35) | 61 (33) | 49 (39) |  |
| *APOE* ε4 carrier, n (%) | 115 (40) | 92 (38) | 71 (38) | 54 (43) | 0.8* |
| Bristol stool score, mean (SD) | 4.0 (1.2) | 4.0 (1.2) | 3.9 (1.2) | 4.0 (1.1) | 0.6^☨^ |
| Race, n (%) |  |  |  |  | 0.4^✝^ |
| White | 257 (90) | 225 (93) | 175 (94) | 120 (94) |  |
| Black or African American | 28 (9.8) | 15 (6.2) | 9 (4.8) | 7 (5.5) |  |
| Other | 2 (0.7) | 2 (0.8) | 2 (1.1) | 0 (0) |  |
| Body mass index, mean (SD) kg/m^2^ | 29 (6.3) | 29 (6.2) | 28 (5.5) | 28 (5.3) | 0.7^☨^ |
| Educational attainment, n (%) |  |  |  |  | 0.8^✝^ |
| High school or less | 36 (13) | 23 (9.5) | 19 (10) | 11 (8.7) |  |
| Some college through Bachelor’s degree | 131 (46) | 110 (45) | 79 (42) | 56 (44) |  |
| Postgraduate degree | 120 (42) | 109 (45) | 88 (47) | 60 (47) |  |
| Fecal SCFA abundance, median normalized |  |  |  |  |  |
| Acetate | 1.0 (0.35) | 1.1 (0.37) | 1.0 (0.36) | 1.1 (0.40) | 0.3^☨^ |
| Propionate | 1.1 (0.62) | 1.1 (0.63) | 1.1 (0.56) | 1.1 (0.65) | 0.8^☨^ |
| Isobutyrate | 1.1 (0.63) | 1.1 (0.67) | 1.1 (0.73) | 1.1 (0.47) | >0.9^☨^ |
| Butyrate | 1.3 (0.96) | 1.3 (0.97) | 1.3 (1.0) | 1.4 (1.0) | 0.7^☨^ |
| Isovalerate | 1.2 (0.84) | 1.2 (0.89) | 1.2 (0.96) | 1.1 (0.56) | 0.2^☨^ |
| Valerate | 1.2 (1.2) | 1.2 (1.3) | 1.2 (1.4) | 1.1 (0.64) | 0.13^☨^ |

*Pearson's Chi-squared test ^✝^Fisher's exact test ^☨^One-way analysis of means (not assuming equal variances). Race was self-reported by participants; races classified as Other include American Indian or Alaska Native (n=1), Asian (n=1), and multiple races (n=1). Abbreviations: A+, amyloid positive status determined using ^11^C-PiB PET, CSF Aβ_42_/Aβ_40_, or plasma pTau_217_; *APOE*, apolipoprotein E; SCFA, short-chain fatty acid; T+, tau positive status determined using ^18^F-MK6240

**Supplementary Table 4. Participant demographics compared across subsets with longitudinal plasma pTau217 or cognitive data.**

| **Characteristic** | **Cognitive data**  **Goal 1**  N = 283 | **Plasma pTau217**  **Goal 1**  N = 219 | ***P* value** |
| --- | --- | --- | --- |
| Clinical diagnosis, n (%) |  |  | 0.002* |
| Cognitively unimpaired | 246 (87) | 208 (95) |  |
| Dementia | 37 (13) | 11 (5.0) |  |
| A+ at fecal collection, n (%) | 65 (27) | 54 (26) | 0.8* |
| Unknown amyloid status, n | 42 | 11 |  |
| T+ at fecal collection, n (%) | 31 (17) | 28 (17) | >0.9* |
| Unknown tau status, n | 99 | 53 |  |
| Age at fecal collection, mean (SD) years | 68 (7.0) | 68 (6.5) | >0.9^☨^ |
| Sex, n (%) |  |  | 0.8* |
| Female | 181 (64) | 138 (63) |  |
| Male | 102 (36) | 81 (37) |  |
| *APOE* ε4 carrier, n (%) | 113 (40) | 78 (36) | 0.3* |
| Bristol stool score, mean (SD) | 4.0 (1.2) | 4.0 (1.2) | >0.9^☨^ |
| Race, n (%) |  |  | 0.3^✝^ |
| White | 254 (90) | 204 (93) |  |
| Black or African American | 27 (9.5) | 13 (5.9) |  |
| Other | 2 (0.7) | 2 (0.9) |  |
| Body mass index, mean (SD) kg/m^2^ | 29 (6.1) | 29 (6.0) | 0.8^☨^ |
| Educational attainment, n (%) |  |  | 0.3^✝^ |
| High school or less | 34 (12) | 19 (8.7) |  |
| Some college through Bachelor’s degree | 129 (46) | 96 (44) |  |
| Postgraduate degree | 120 (42) | 104 (47) |  |
| Fecal SCFA abundance, median normalized |  |  |  |
| Acetate | 1.0 (0.35) | 1.0 (0.35) | 0.6^☨^ |
| Propionate | 1.1 (0.64) | 1.1 (0.57) | 0.2^☨^ |
| Isobutyrate | 1.1 (0.64) | 1.1 (0.68) | >0.9^☨^ |
| Butyrate | 1.3 (0.96) | 1.2 (0.92) | 0.6^☨^ |
| Isovalerate | 1.2 (0.86) | 1.2 (0.90) | 0.8^☨^ |
| Valerate | 1.3 (1.3) | 1.2 (1.3) | 0.7^☨^ |

*Pearson's Chi-squared test ^✝^Fisher's exact test ^☨^One-way analysis of means (not assuming equal variances). Race was self-reported by participants; races classified as Other include American Indian or Alaska Native (n=1), Asian (n=1), and multiple races (n=1). Abbreviations: A+, amyloid positive status determined using ^11^C-PiB PET, CSF Aβ_42_/Aβ_40_, or plasma pTau_217_; *APOE*, apolipoprotein E; SCFA, short-chain fatty acid; T+, tau positive status determined using ^18^F-MK6240

**Supplementary Tables 5-24 can be found in the attached excel file.**
